# State of play in individual participant data meta-analyses of randomised trials: Systematic review and consensus-based recommendations

**DOI:** 10.64898/2026.02.03.26345481

**Authors:** Anna Lene Seidler, Jannik Aagerup, Lily S Nicholson, Kylie E Hunter, Ram Bajpai, Daniel G Hamilton, Thomas Love, Nadine Marlin, David Nguyen, Richard D Riley, Larysa HM Rydzewska, Mark Simmonds, Lesley A Stewart, Wilson Tam, Jayne F Tierney, Rui Wang, Alain Amstutz, Matthias Briel, Sarah Burdett, Joie Ensor, Miriam Hattle, Sol Libesman, Yiwen Liu, Stefan Schandelmaier, Lianne K Siegel, Kym IE Snell, James Sotiropoulos, Claire L Vale, Ian R White, Jonathan G Williams, Peter J Godolphin

## Abstract

**Background:** Individual participant data (IPD) meta-analyses obtain, harmonise and synthesise the raw individual-level data from multiple studies, and are increasingly important in an era of data sharing and personalised medicine to inform clinical practice and policy.

**Objectives:** (1) Describe the landscape of IPD meta-analysis of randomised trials over time;

(2) establish current practice in design, conduct, analysis and reporting for pairwise IPD meta-analysis; and (3) derive recommendations to improve the conduct of and methods for future IPD meta-analyses.

**Design:** Part 1: systematic review of all published IPD meta-analyses of randomised trials; Part 2: in-depth review of current methodological practice for pairwise IPD meta-analysis; and Part 3: adapted nominal group technique to derive consensus recommendations for IPD meta-analysis authors, educators and methodologists.

**Data sources:** MEDLINE, Embase, and the Cochrane Database of Systematic Reviews (via the Ovid interface).

**Eligibility criteria:** Part 1: all IPD meta-analyses of randomised trials published before February 2024, evaluating intervention effects and based on a systematic search. Part 2: all pairwise IPD meta-analyses from part 1 published between February 2022 and February 2024. Part 3: Selected panel of experienced IPD meta-analysis authors and/or methodologists.

**Results:** Part 1: We identified 605 eligible IPD meta-analyses published between 1991 and 2024. The number of IPD meta-analyses published per year increased over time until 2019 but has since plateaued to about 60 per year. The most common clinical areas studied were cardiovascular disease (n=113, 19%) and cancer (n=110, 18%). The proportion of IPD meta-analyses published with Cochrane decreased over time from 16% (n=31/196) before 2015 to 3% (n=5/196) between 2021-2024. Part 2: 100 recent pairwise IPD meta-analyses were included in the in-depth review. Most cited PRISMA-IPD (68, 68%) and conducted risk of bias assessments (n=82, 82%), with just under half carrying out subgroup analyses not at risk of aggregation bias (n=36/85, 41%). However, only 33% (n=33) and 29% (n=29) respectively provided a protocol or statistical analysis plan, and only 7% (n=6/82) reported using IPD to inform risk of bias assessments. Part 3: 24 experts participated in a consensus workshop. Key recommendations for improved IPD meta-analyses focused on transparency (prospective registration; published protocols and statistical analysis plans) and maximising value (searching trial registries; obtaining IPD for unpublished evidence; using IPD to address missing data and risk of bias). Methodologists and educators should strengthen dissemination of methods and support capacity building across clinical fields and geographical areas.

**Conclusions:** The application and methodological quality of IPD meta-analyses of randomised trials has increased in the last decade, but shortcomings remain. Implementing our consensus-based recommendations will ensure future IPD meta-analyses generate better evidence for clinical decision making.

**Study registration:** Open Science Framework (1)

**Summary boxes:** *What is already known on this topic:*

- IPD meta-analyses of randomised trials are regularly used to inform clinical policy and practice.
- They can provide better quality data and enable more thorough and robust analyses than standard aggregate data meta-analyses, but are resource-intensive and can be challenging to conduct, leading to variable methodological quality
- Previous studies that evaluated the conduct of IPD meta-analyses pre-date several major developments, such as the introduction of the PRISMA-IPD reporting guideline.

*What this study adds:*

- This is the most comprehensive assessment of IPD meta-analyses of randomised trials to date (605 studies), showing an increase in publications over time followed by a recent plateau.
- The conduct of IPD meta-analysis has improved in recent years including increased use of prospective registration, assessment of risk of bias, appropriate analyses of patient subgroup effects and citing the PRISMA-IPD statement.
- Many shortcomings remain including (i) insufficient pre-specification of methods such as outcomes and analyses, (ii) sub-standard transparency (including publication of protocols, statistical analysis plans and reporting of analyses), and (iii) failure to gain maximum value of IPD (i.e. include unpublished trials, use the IPD to inform risk of bias and trustworthiness assessments, and address missing data appropriately); expert consensus recommendations are provided for how to address these gaps.

## Introduction

Systematic reviews and meta-analyses inform policy and practice but are typically limited to the synthesis of aggregate data (i.e. study-level summary information) extracted from publications or other records of eligible primary studies. Individual participant data (IPD) meta-analysis is an alternative approach which involves collection, harmonisation and synthesis of the raw individual-level data for eligible studies (2).

IPD meta-analysis has been called the ‘gold standard’ due to the many advantages it provides, including higher data availability and the use of more appropriate and consistent analysis methods (2). IPD meta-analysis may provide more reliable results that better inform clinical practice and policy (2,3), especially for personalised healthcare (precision medicine) where healthcare decisions are tailored to patient characteristics. Yet historically, IPD meta-analyses have not always followed best practice methodology on design, conduct, analysis and reporting, undermining these advantages (4,5).

To improve the quality of IPD meta-analysis, an accurate and up to date understanding of current practice in the field is needed. This can enable identification of where new methods, guidance and reporting standards need to be developed and implemented. However, key previous reviews of the features and methods of IPD meta-analyses are now over a decade old (4–7). These reviews identified major limitations in the conduct and reporting of IPD meta-analyses. For example, a review of studies from 2008 to 2014 identified that few assessed risk of bias or reported data retrieval rates, data management processes or how data were checked (5). More recently, a review focused on the methodological quality of IPD meta-analysis published between 1991 and 2019 but did not cover items around conduct and reporting, or analytical strategies (8). Since these reviews were published, much has changed in the IPD meta-analysis field, including new best practice methodology (e.g. subgroup analyses not subject to aggregation bias (9,10)), the introduction of data sharing policies and requirements (11–13), reporting guidelines (14) and assessment of IPD for integrity issues (15).

In this article, we present the most comprehensive and up-to-date overview of developments and changes in IPD meta-analysis practice over time and outline the current state of play in IPD meta-analyses of randomised trials. We provide consensus recommendations from leading methodological experts on best practice for planning, conducting and reporting of future IPD meta-analyses.

## Methods

Our study was guided by a prospectively registered protocol (1). It was divided into three parts: (1) a systematic review of all IPD meta-analyses of randomised trials evaluating intervention effects, aiming to describe the landscape of IPD meta-analysis over time; (2) an in-depth review of current practice (2022-2024) in conduct, analysis and reporting of pairwise IPD meta-analyses; (3) a consensus process to derive recommendations to inform and improve the conduct and reporting of future IPD meta-analyses of randomised trials.

Parts 1 and 2 were reported in accordance with the PRISMA 2020 statement where applicable (16) (Supplementary File 1) and part 3 using the ACCORD guideline (17) (Supplementary File 2).

### Parts 1 & 2: Systematic review and in-depth review

#### Eligibility, information sources and search strategy

##### Part 1: Systematic review of the landscape of IPD meta-analysis

We systematically searched MEDLINE, Embase, and the Cochrane Database of Systematic Reviews (all via the Ovid interface) for published IPD meta-analyses of randomised trials evaluating the effect of interventions. We only included IPD meta-analyses based on a systematic search (where systematic literature searches were conducted in at least 2 databases). Both pairwise IPD meta-analyses (i.e. comparing two interventions) and network IPD meta-analyses (i.e. comparing more than two interventions) were included in part 1. IPD meta-analyses using only single arms from randomised trials and studies focusing on prevalence, prognosis, surrogacy, economic evaluations and diagnostic test accuracy were excluded. Protocol papers and publications not in English were excluded. This search was previously performed up to September 2019 in a systematic review by Wang et al. (8);we updated their search to 26 February 2024 (see Supplementary File 3 for our full search strategy).

##### Part 2: In-depth review of current practice

For part 2, eligible studies were all pairwise IPD meta-analyses identified in part 1 that were published across a two-year period (26 February 2022 to 26 February 2024). IPD meta-analyses that included a network meta-analysis were excluded from part 2.

#### Screening and data extraction

Titles and abstracts were initially independently screened in duplicate by two reviewers, followed by full-text reports, in Covidence systematic review software (18). Discrepancies were resolved either by a third reviewer or through consensus. All data extraction fields and definitions are available in the published protocol (1), and final data extraction forms including definitions of each variable can be found in the Supplementary material (Supplementary Files 4 and 5). Authors of included IPD meta-analyses were not involved in the screening or extraction of their own studies. Our approach to data extraction differed for parts 1 and 2:

##### Part 1: Systematic review of the landscape of IPD meta-analysis

We extracted basic characteristics of the IPD meta-analyses, following a similar approach to Wang et al. (8) to enable comparison. We calculated the impact factor and journal quartile using a consistent year for each study (2025). Medical area was coded according to the International Classification of Diseases 11th Revision (19). All items were extracted in duplicate with disagreements resolved by consensus.

##### Part 2: In-depth review of current practice

We developed a data extraction form to capture information on data retrieval, reporting, conduct, and analysis approaches (Supplementary File 5). Data fields were guided by items in PRISMA-IPD (14), previous IPD methods projects, (3,5,7,20,21) and input from our assembled team of IPD methodologists, statisticians, and evidence synthesis experts. This was tested on five eligible studies and subsequently refined.

For each pair of reviewers, data extraction was initially performed independently in duplicate for the first 40% of the eligible studies, with disagreements resolved by consensus with a third reviewer. Once agreement was near perfect, we switched to single extraction for the remaining studies. Near perfect was defined as no disagreements or only minor non-consequential disagreements. This deviated from the protocol, which had specified a kappa threshold, but the number of data items made calculation of kappa for each reviewer pair and item infeasible.

#### Analysis

All analyses were performed in Stata version 18.5 (StataCorp, Texas, USA) or later and were descriptive, with frequency calculated (%) for categorical data and median, interquartile range (IQR) and range for continuous data.

### Part 3: Consensus process to derive recommendations

#### Selection of panel members

The steering committee consisted of two experienced IPD meta-analysis methodologists (PJG and ALS) and three early-career researchers (JA, LSN, DN). The steering committee assembled a panel of experts in IPD meta-analysis to come together to interpret our findings, derive recommendations for future IPD meta-analyses and identify untapped opportunities in the field. Panel members were selected and invited by the lead and senior author, with panel members offered the opportunity to suggest additional candidates.

Criteria for inclusion in the panel were: lead or senior author on a previous IPD meta-analysis or on an important methods article, book or book chapter on IPD meta-analysis, or having a leading role in an IPD methods group (including Cochrane IPD Meta-analysis Methods group and SMART-IPD methods group). We aimed to include participants with diverse expertise (e.g. statisticians, physicians, systematic reviewers, information specialists, methodologists, health technology assessors), as well as variation in medical subject specialty, career stage and geographical location.

#### Assessing consensus

The consensus process was informed by the results of our systematic review of IPD meta-analyses and in-depth review (part 1 and 2 of this study), which were collated into a detailed report and worksheet for the consensus meeting and distributed to panel members in advance (see Supplementary File 6).

We then conducted four consensus workshops, with a maximum of eight members per workshop group to enable every panel member to contribute meaningfully, while also including a wide range of perspectives and backgrounds in each group. The workshops were scheduled to last two hours, with each member invited to only one workshop.

To assess consensus, we used a modified nominal group technique (22). This comprised four stages: 1. Silent individual ideas generation; 2. Round robin collation of ideas; 3.

Clarification of ideas; and 4. Agreeing on key recommendations. During the silent individual ideas generation and round robin, panel members were asked which of the results were most noteworthy and whether there were key take aways, lessons or recommendations. As we wanted to include all recommendations the groups agreed on, no formal ranking of recommendations was deemed necessary. Thus, instead of conducting a formal vote, each consensus workshop group reached agreement on the key recommendations through discussion.

First, we aimed to reach consensus on the interpretation of results from the systematic review and in-depth review (parts 1 and 2), which we organised into six sections (see Supplementary File 6). Participants were shown one section of results at a time (via a table and/or figure). We used the first three stages of the nominal group technique to establish key interpretation points from each section. Subsequently, each group went through the full nominal group technique for two specific questions when considering the totality of the results: (1) “What do these results tell us about the current state of play in IPD meta-analysis?”, and (2): “Where do we need to develop new guidance or provide better access to existing methods?”.

The process was piloted by the steering committee and subsequently refined. Each discussion group was facilitated by either ALS or PJG, with one additional steering committee group member taking detailed minutes. The consensus from each of the four groups was synthesised into overall themes by ALS following a thematic analysis approach (23) ensuring that recommendations from multiple meetings would only appear once in the overall summary. These themes were then cross-checked against the minutes by two additional steering committee members (PJG and JA) to capture any omitted themes.

### Patient and Public Involvement

There was no patient or public involvement. This study was co-designed with important stakeholders in the field of IPD meta-analysis, including methods developers, reporting guideline developers, methods group leads and authors of large IPD meta-analyses; to ensure relevance to the target group (‘consumers’) of this research.

## Results

### Part 1: Systematic review of the landscape of IPD meta-analysis

For the updated search (September 2019 to February 2024), we screened titles and abstracts of 4588 studies and full texts of 848 studies, identifying 282 eligible IPD meta-analyses. Additionally, we included all 323 IPD meta-analyses identified in the previous review up to 2019 (8). Thus, we identified a total of 605 IPD meta-analyses of randomised trials evaluating interventions with a systematic search for inclusion in part 1 (see Supplementary Figure 1).The full dataset for parts 1 and 2 with references for all studies is available on Open Science Framework (1).

The 605 IPD meta-analyses were published between 1991 and 2024. The number published per year increased over time, up to a peak of 73 in 2019; since then numbers have plateaued at approximately 60 per year (Figure 1). The main clinical areas studied were diseases of the circulatory system (n=113/605, 19%), neoplasms (n=110/605, 18%), and mental, behavioural or neurodevelopmental disorders (n=75/605, 12%) (Table 1). The majority of IPD meta-analyses had corresponding authors based in Europe (n=424/605, 70%). Across the IPD meta-analyses, the median number of trials that provided IPD was 7 (IQR 4 to 14), including a median total sample size across trials of 2127 participants (IQR 810 to 5948). The number of IPD meta-analyses that were Cochrane reviews fell from 16% (n=31/196) for studies published in or before 2015 to 3% (n=5/196) for studies published between 2021 and 2024. For IPD meta-analyses published after September 2019, PROSPERO registration increased from 44% (2019-2020) to 65% (2021-2024).

**Figure 1:**
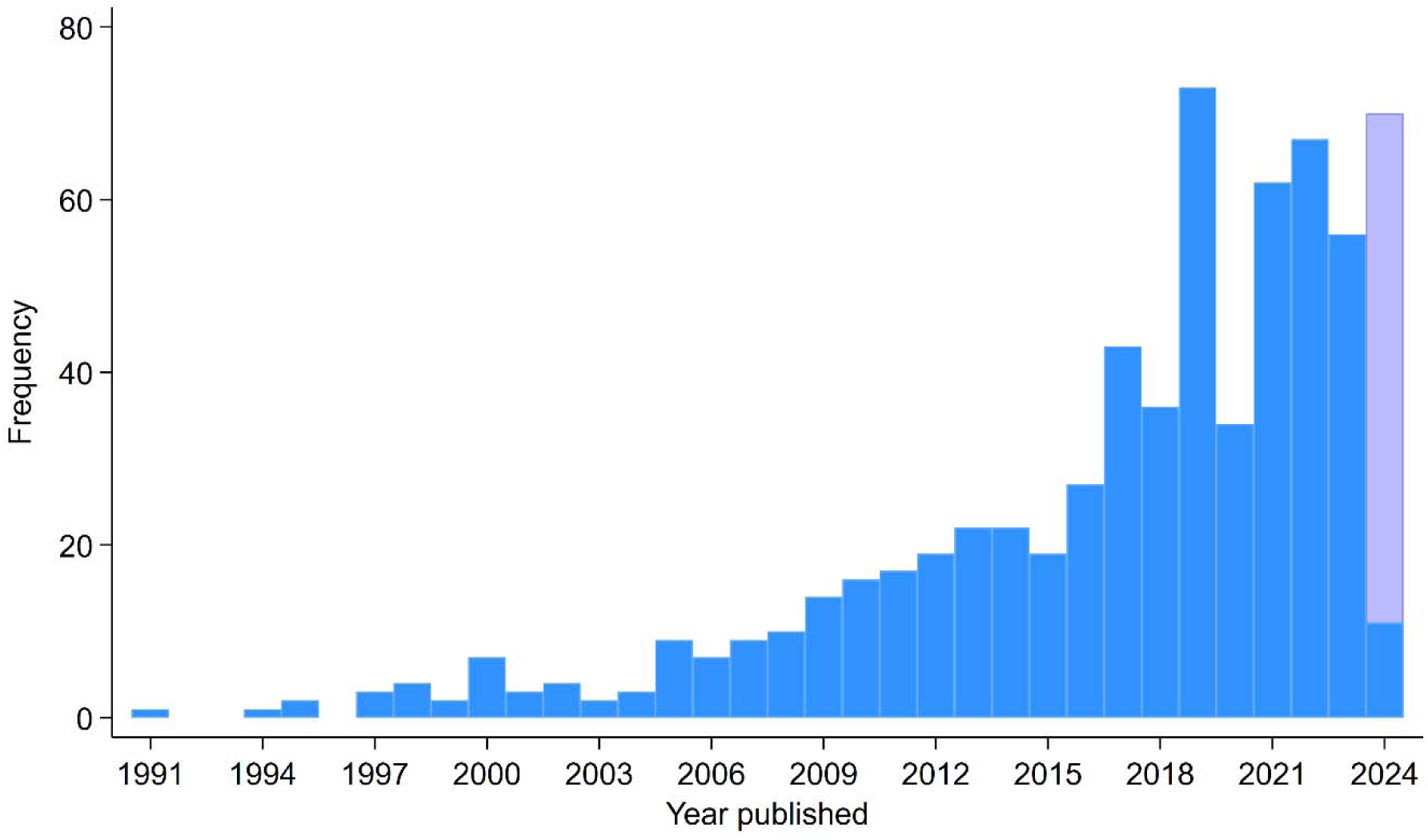
Number of individual participant data meta-analyses of randomised trials that were part of a systematic review over time. The 2024 data are restricted up to 26^th^ February 2024 – the translucent bar shows this extrapolated to the end of 2024 by assuming that the number of published individual participant data meta-analyses continues at the same rate as between 01^st^ January 2024 – 26^th^ February 2024.

**Table 1:**
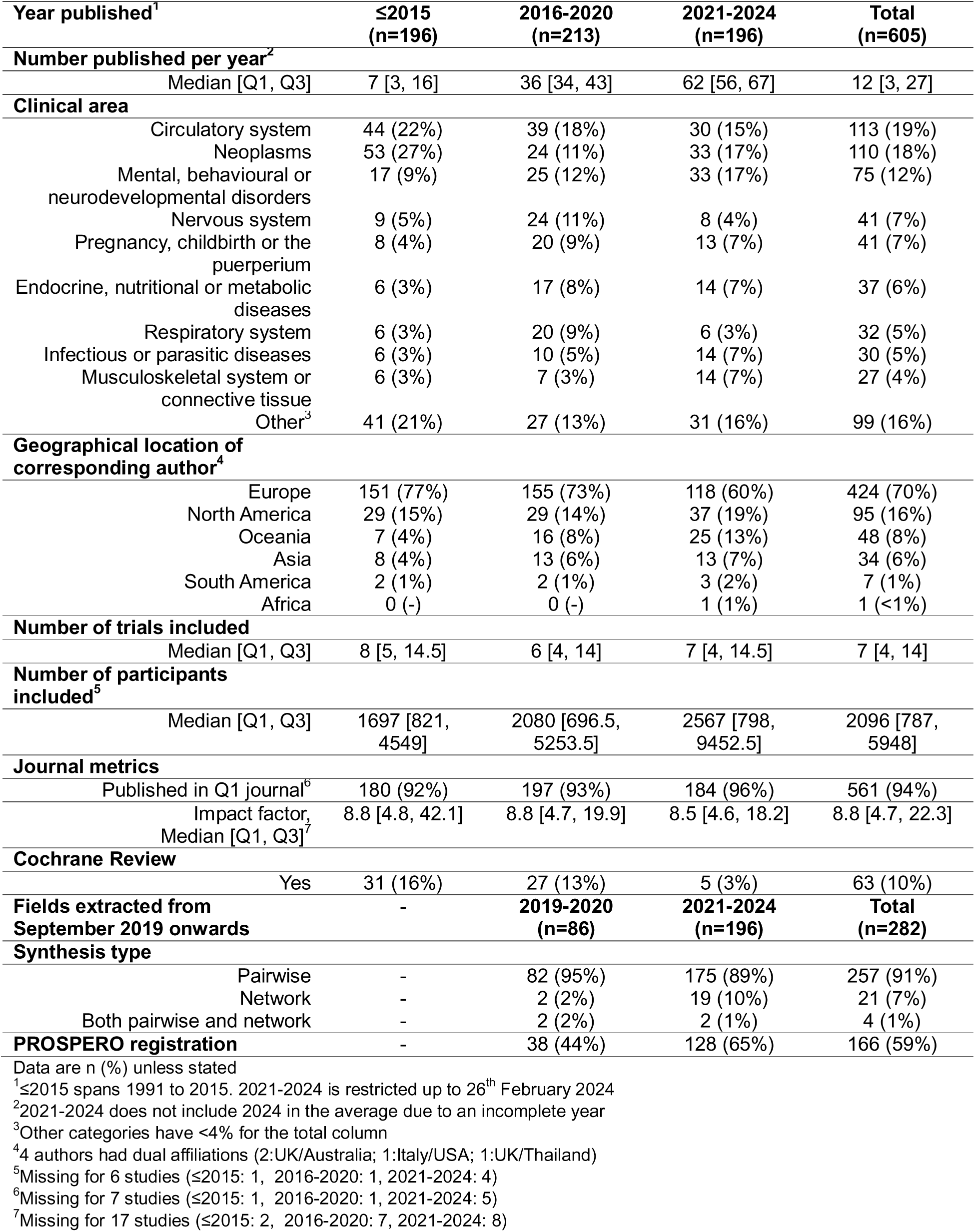
General characteristics of the 605 IPD meta-analyses of randomised trials identified in the systematic review

| Year published<sup>1</sup> | ≤2015<br>(n=196) | 2016-2020<br>(n=213) | 2021-2024<br>(n=196) | Total<br>(n=605) |
| --- | --- | --- | --- | --- |
| **Number published per year<sup>2</sup>** |  |  |  |  |
| Median [Q1, Q3] | 7 [3, 16] | 36 [34, 43] | 62 [56, 67] | 12 [3, 27] |
| **Clinical area** |  |  |  |  |
| Circulatory system | 44 (22%) | 39 (18%) | 30 (15%) | 113 (19%) |
| Neoplasms | 53 (27%) | 24 (11%) | 33 (17%) | 110 (18%) |
| Mental, behavioural or neurodevelopmental disorders | 17 (9%) | 25 (12%) | 33 (17%) | 75 (12%) |
| Nervous system | 9 (5%) | 24 (11%) | 8 (4%) | 41 (7%) |
| Pregnancy, childbirth or the puerperium | 8 (4%) | 20 (9%) | 13 (7%) | 41 (7%) |
| Endocrine, nutritional or metabolic diseases | 6 (3%) | 17 (8%) | 14 (7%) | 37 (6%) |
| Respiratory system | 6 (3%) | 20 (9%) | 6 (3%) | 32 (5%) |
| Infectious or parasitic diseases | 6 (3%) | 10 (5%) | 14 (7%) | 30 (5%) |
| Musculoskeletal system or connective tissue | 6 (3%) | 7 (3%) | 14 (7%) | 27 (4%) |
| Other<sup>3</sup> | 41 (21%) | 27 (13%) | 31 (16%) | 99 (16%) |
| **Geographical location of corresponding author<sup>4</sup>** |  |  |  |  |
| Europe | 151 (77%) | 155 (73%) | 118 (60%) | 424 (70%) |
| North America | 29 (15%) | 29 (14%) | 37 (19%) | 95 (16%) |
| Oceania | 7 (4%) | 16 (8%) | 25 (13%) | 48 (8%) |
| Asia | 8 (4%) | 13 (6%) | 13 (7%) | 34 (6%) |
| South America | 2 (1%) | 2 (1%) | 3 (2%) | 7 (1%) |
| Africa | 0 (-) | 0 (-) | 1 (1%) | 1 (<1%) |
| **Number of trials included** |  |  |  |  |
| Median [Q1, Q3] | 8 [5, 14.5] | 6 [4, 14] | 7 [4, 14.5] | 7 [4, 14] |
| **Number of participants included<sup>5</sup>** |  |  |  |  |
| Median [Q1, Q3] | 1697 [821, 4549] | 2080 [696.5, 5253.5] | 2567 [798, 9452.5] | 2096 [787, 5948] |
| **Journal metrics** |  |  |  |  |
| Published in Q1 journal<sup>6</sup> | 180 (92%) | 197 (93%) | 184 (96%) | 561 (94%) |
| Impact factor, Median [Q1, Q3]<sup>7</sup> | 8.8 [4.8, 42.1] | 8.8 [4.7, 19.9] | 8.5 [4.6, 18.2] | 8.8 [4.7, 22.3] |
| **Cochrane Review** |  |  |  |  |
| Yes | 31 (16%) | 27 (13%) | 5 (3%) | 63 (10%) |
| **Fields extracted from September 2019 onwards** | - | **2019-2020**<br>**(n=86)** | **2021-2024**<br>**(n=196)** | **Total**<br>**(n=282)** |
| **Synthesis type** |  |  |  |  |
| Pairwise | - | 82 (95%) | 175 (89%) | 257 (91%) |
| Network | - | 2 (2%) | 19 (10%) | 21 (7%) |
| Both pairwise and network | - | 2 (2%) | 2 (1%) | 4 (1%) |
| **PROSPERO registration** | - | 38 (44%) | 128 (65%) | 166 (59%) |

Data are n (%) unless stated
<sup>1</sup>≤2015 spans 1991 to 2015. 2021-2024 is restricted up to 26<sup>th</sup> February 2024
<sup>2</sup>2021-2024 does not include 2024 in the average due to an incomplete year
<sup>3</sup>Other categories have <4% for the total column
<sup>4</sup>4 authors had dual affiliations (2:UK/Australia; 1:Italy/USA; 1:UK/Thailand)
<sup>5</sup>Missing for 6 studies (≤2015: 1, 2016-2020: 1, 2021-2024: 4)
<sup>6</sup>Missing for 7 studies (≤2015: 1, 2016-2020: 1, 2021-2024: 5)
<sup>7</sup>Missing for 17 studies (≤2015: 2, 2016-2020: 7, 2021-2024: 8)

### Part 2: In-depth review of current practice

Of the IPD meta-analyses identified in the systematic review, 101 were published between 26 February 2022 and 26 February 2024. After excluding a single network IPD meta-analysis, 100 pairwise IPD meta-analyses were included in the in-depth review.

#### General characteristics, data sources and retrieval

The 100 recent IPD meta-analyses were spread across a similar set of clinical areas with the same top three as in the total sample, albeit in a different order (Supplementary Table 1).

Only 3 were Cochrane reviews and 13 were either entirely prospective (n=9) or partially prospective (i.e. a “nested prospective” meta-analysis (24) (n=4)) (Figure 2)(24). Most IPD meta-analyses were funded through public or philanthropic sources (n=70); only 13 IPD meta-analyses reported industry involvement. Most IPD meta-analyses were conducted in high-income, Western countries (58 Europe, 18 Australia, 14 USA, Figure 2).

**Figure 2.**
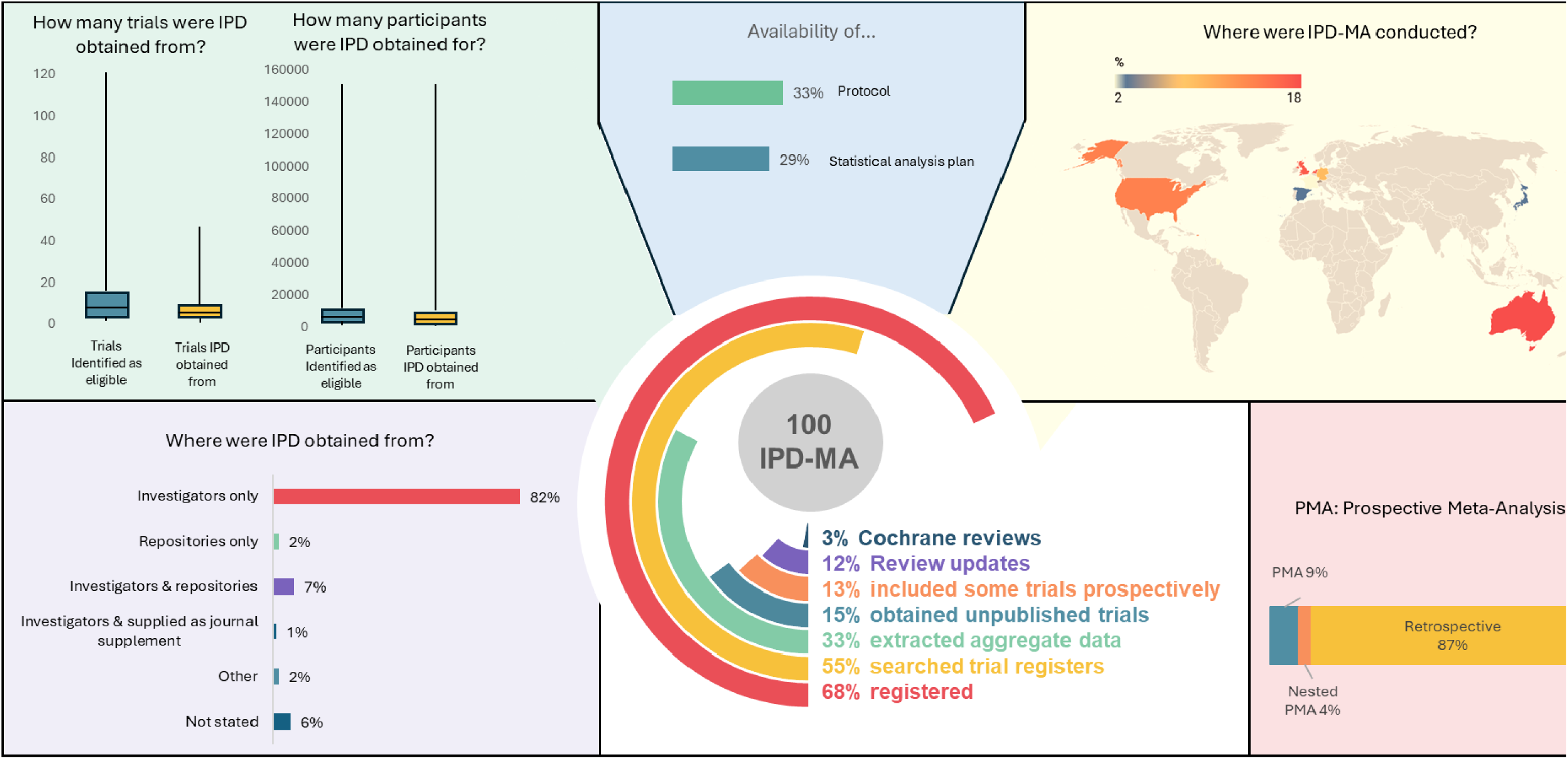
General characteristics, data sources and retrieval for 100 IPD meta-analyses published between February 2022 and February 2024

In the subset of 100 current IPD meta-analyses, the median number of included trials was 9 (IQR 4 to 17), and the median number of trials for which IPD were retrieved was 6 (IQR 4 to 10). The median trial IPD retrieval rate was 85.7%, with 41 of 99 (41%, 1 unclear) IPD meta-analyses obtaining IPD from all included trials (Figure 2, Supplementary Table 2). 83 (83%) IPD meta-analyses reported the number of included participants. From these, the median reported participant IPD retrieval rate was 99.2% (IQR 75.6% to 100%), with 39 of 83 IPD meta-analyses reporting that information (47%) obtaining data on all eligible participants.

The majority of IPD was obtained through contact with investigators, with only 9 (9%) reporting that they used data repositories to access all or part of the IPD. Aggregate data from randomised trials for which IPD could not be retrieved was extracted in a third of the included IPD meta-analyses (33%, n=33). Clinical trial register searches were reported by 55 (55%) of the IPD meta-analyses; 15 (15%) IPD meta-analyses obtained IPD from unpublished trials.

#### Design, conduct and reporting

A clear rationale for conducting the IPD meta-analysis was given in 62 (62%) of cases. The main reported reasons were investigation of subgroup effects/effect modification (n=45/62, 73%) and ability to better estimate an overall treatment effect (n=42/62, 68%) (Figure 3, Supplementary Table 3). About two-thirds of IPD meta-analyses (n=68, 68%) were registered on PROSPERO, but a protocol was only identified for one third (n=33, 33%), and a separate statistical analysis plan in less than one third (n=29, 29%). The majority of studies (n=66, 66%) stated that they followed PRISMA-IPD (14), but 17 (17%) IPD meta-analyses did not refer to any reporting guideline.

**Figure 3.**
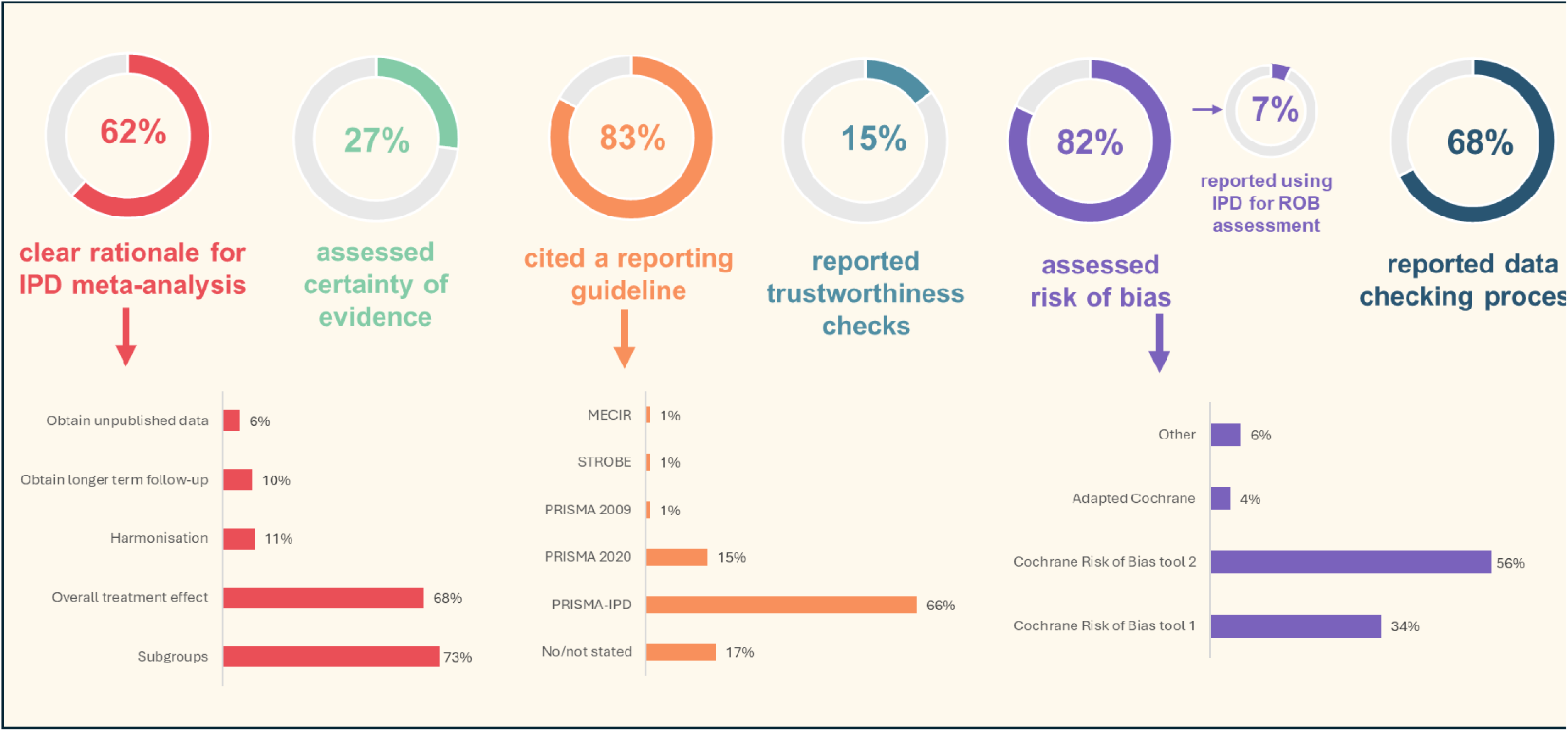
Design, conduct and reporting for 100 IPD meta-analyses published between February 2022 and February 2024

Risk of bias assessments were reported in most IPD meta-analyses (82%), most frequently using the Cochrane Risk of Bias 2 tool (n=46/82, 56%) (Figure 3, Supplementary Table 4). IPD was rarely reported as being used itself to inform risk of bias assessments in addition to published reports (n=6/82, 7%). A data checking process for the IPD obtained was reported in around two thirds of IPD meta-analyses (n=68, 68%). However, the details provided were often insufficient to fully understand or replicate these checks. Assessments of the integrity of included trials (i.e., trustworthiness checks) were reported in 15 (15%) IPD meta-analyses, and certainty of evidence was assessed in 27 (27%). Of these, 26 (96%) reported using GRADE (25).

#### Statistical analysis strategies

An estimand (i.e. the specific treatment effect of interest (26)) was only reported in one IPD meta-analysis (1%), and only 4 (4%) described a sample size or power calculation (Figure 4, Supplementary Table 5). Aggregate data (obtained for randomised trials for which IPD was not available) was rarely combined with the IPD as part of the main analysis (n=9, 9%) but was more commonly included in a sensitivity analysis (n=16, 16%). A one-stage analysis (where the IPD from all studies are meta-analysed in a single step) was the most common statistical modelling approach for the primary analysis (n=52; 52%). Half used a random-effects model as their main analysis model to allow for between-study heterogeneity (n=50, 50%), 30 (30%) used a common/fixed-effect model as their main analysis model;, the others used both without specifying a main model (n=7, 7%), or were unclear (n=13, 13%). The main analysis model was explicitly adjusted for prognostic factors in half of the IPD meta-analyses; just over half (n=55, 55%) reported using an approach to address missing data.

**Figure 4:**
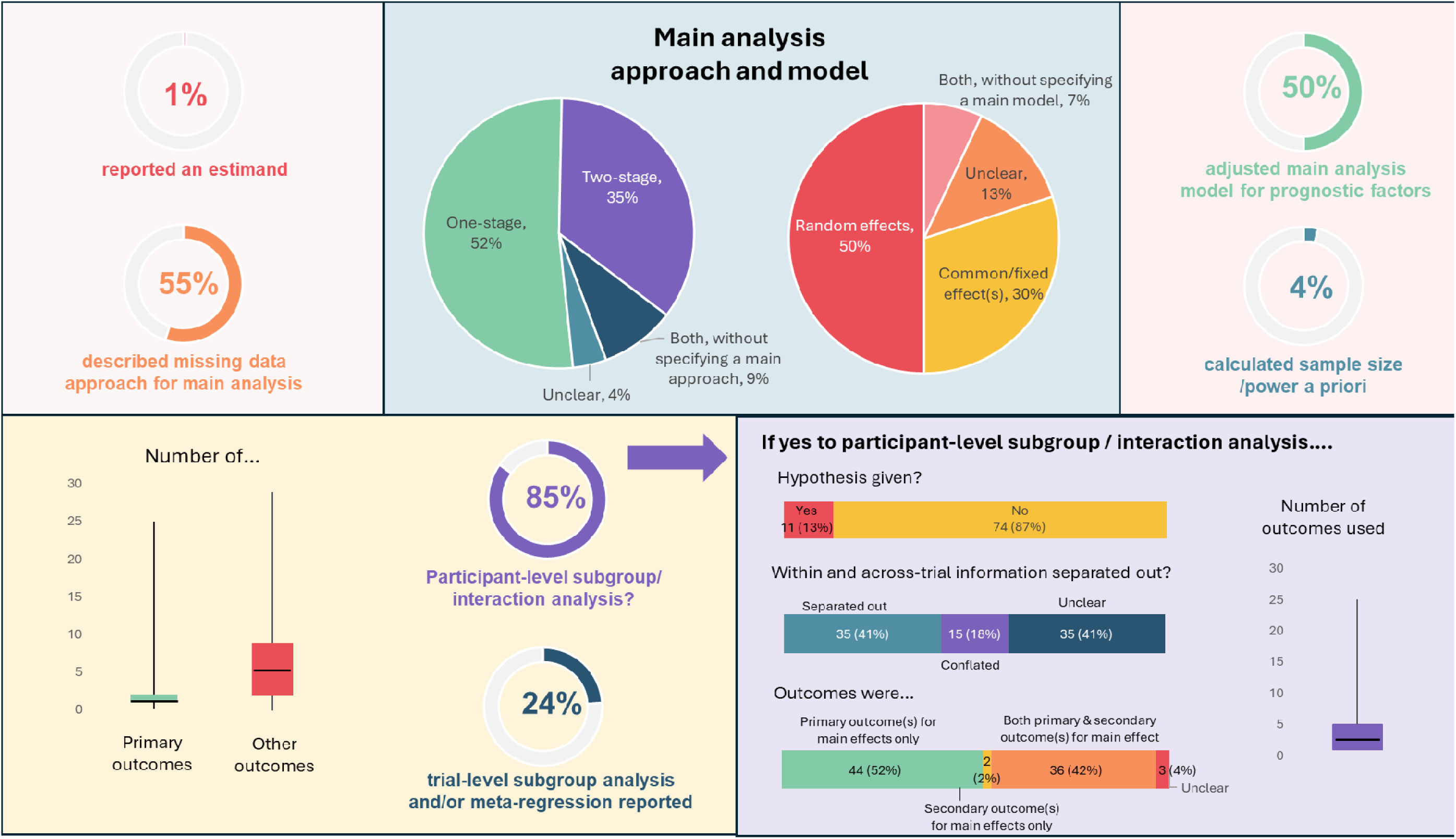
Statistical analysis strategies for 100 IPD meta-analyses published between February 2022 and February 2024

The vast majority (n=85, 85%) of IPD meta-analyses reported conducting at least one participant level subgroup/interaction analysis (Figure 4, Supplementary Table 6). Of those, hypotheses around directionality of the subgroup effect were stated in 11/85 (13%), and a median of 2 (IQR 1 to 5) outcomes, most commonly the IPD meta-analysis primary outcome, were used. For participant-level analyses (i.e. models examining individual-level variability), 35/85 (41%) separated out within- and across-trial information (as is recommended (9,27)), just under a fifth did not separate these out (15/85, 18%) and the remaining 41% (n=35) were unclear. Around a quarter of IPD meta-analyses (n=24, 24%) reported conducting trial-level subgroup analysis or meta-regression, usually in addition to main effect analyses.

### Part 3: Consensus process to derive recommendations

The consensus workshops were held online between 2 and 3 April 2025. We invited 31 experts in IPD meta-analysis to participate in the consensus process; 24 agreed and joined one of four discussion groups (each comprising 4-8 participants). One additional expert was unable to attend but provided written feedback on the results. All participating consensus panel members worked at an academic institution and had expertise in the following areas: undertaking IPD meta-analysis (n=24, 100%), developing IPD meta-analysis methodology (n=19, 79%), systematic reviews (n=19, 79%), statistics (n=11, 46%), clinical trials (n=9, 38%), guideline development (n=5, 21%), clinical medicine (n=4, 17%) and health technology assessment (n=3, 13%). Thirteen panel members had their primary affiliation in the UK, 6 in Australia, 3 in Switzerland, 1 in Singapore and 1 in the USA. The steering committee were also all based at an academic institution, with 2 having their primary affiliation in Germany, 2 in the UK and 1 in Australia. Further characteristics of the panel members can be found in Supplementary Table 7. Panel members were invited to contribute to the manuscript as co-authors.

#### Interpretation of the current state of play of IPD meta-analyses of randomised trials

Table 2 summarises the main themes identified in the consensus process on the landscape and current state of play of IPD meta-analysis (parts 1 and 2).

**Table 2:**
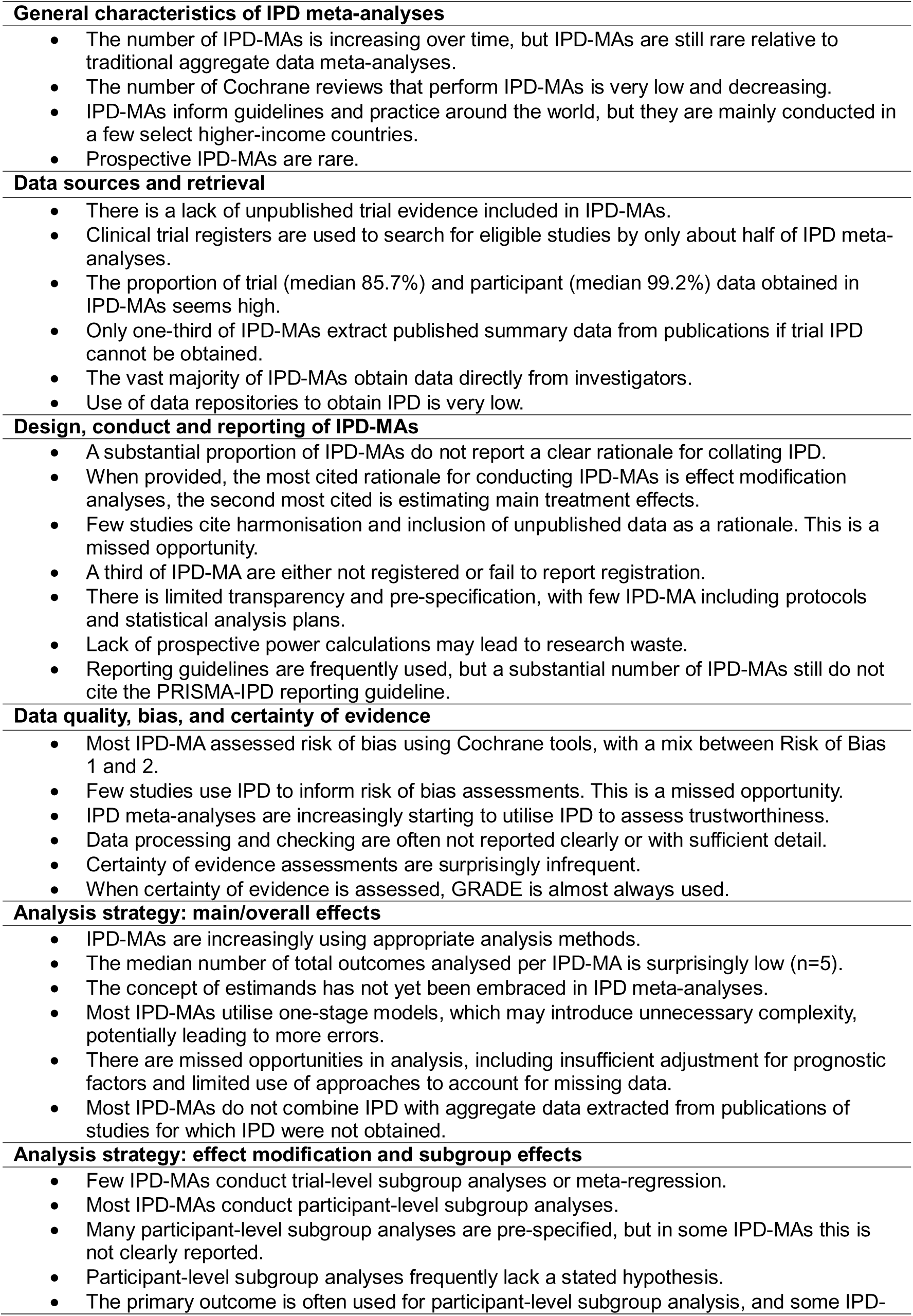
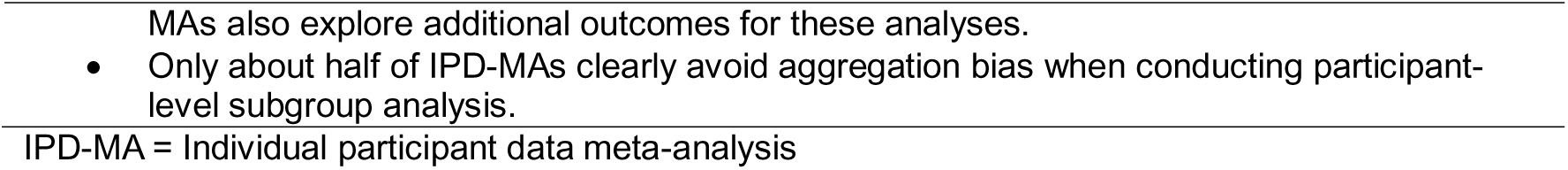
Main themes from the consensus meeting on the landscape and current state of play of individual participant data meta-analyses of randomised trials

The consensus process highlighted that the conduct and reporting of IPD meta-analyses has improved in several respects compared with reviews from 10–20 years ago (4–6). In particular, most recent IPD meta-analyses now cite reporting guidance (often PRISMA-IPD (14), report some level of data checking, with risk of bias assessments now commonplace, and a larger proportion use methods to assess participant-level effect modifiers that do not introduce aggregation bias. However, several areas for improvement were identified, including a lack of transparency and pre-specification (e.g. lack of protocols, statistical analysis plans and available analysis code), major gaps in reporting across all aspects of IPD meta-analysis, underuse of clinical trial registries and unpublished evidence, inconsistent approaches to missing data, limited use of IPD to inform risk of bias assessments and slow and bureaucratic data sharing processes of primary studies making IPD meta-analyses more difficult to conduct, with data repositories rarely used. Further, they noted that the number of IPD meta-analyses has plateaued, with the majority being led by researchers in a few countries and across limited clinical areas.

#### Recommendations, areas for improvement, and future developments

Table 3 summarises the panel’s consensus recommendations for how to address the identified limitations for IPD meta-analysis authors, methodologists and educators. For authors, recommendations focus on making IPD meta-analyses more transparent, reproducible and informative. This includes prospective registration and publication of a full protocol, ensuring that these are sufficiently detailed to be meaningful (i.e. specifying outcomes, analysis populations, main models, and key subgroup hypotheses – ideally including directionality), publication of a statistical analysis plan and making analysis code available alongside clear descriptions of statistical models. To improve reporting, the panel emphasised going beyond simply citing PRISMA-IPD, by explicitly reporting how the guidelines were followed, including key conduct decisions and checks (including how data were requested and processed; what data checks were undertaken and what issues were found/resolved; whether and how IPD informed risk of bias assessments), and clearly reporting statistical analysis decisions and effect modification methods (i.e. what estimands/outcomes were used, whether subgroup analyses were pre-specified; whether within- and across-trial information were separated).

**Table 3:**
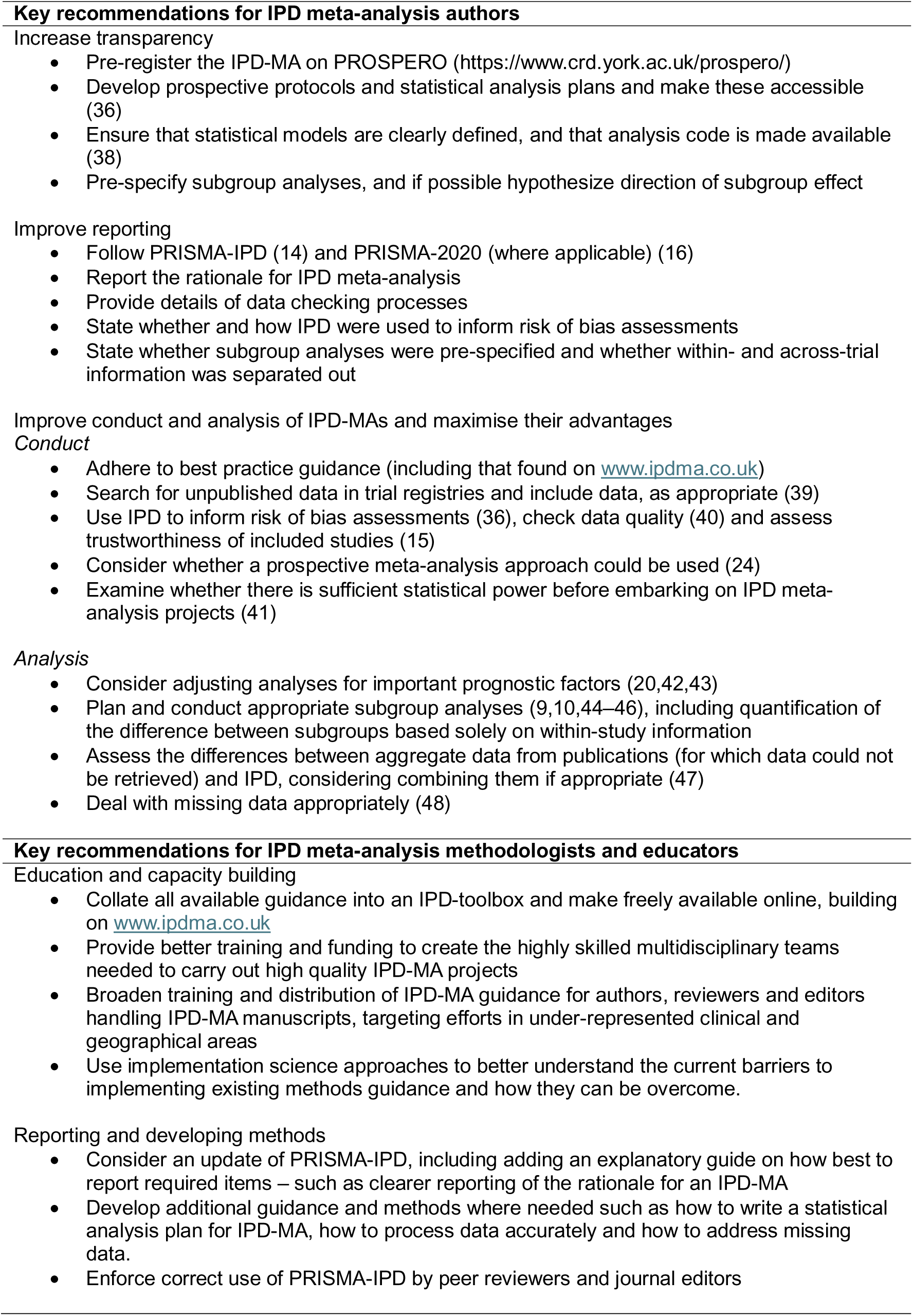
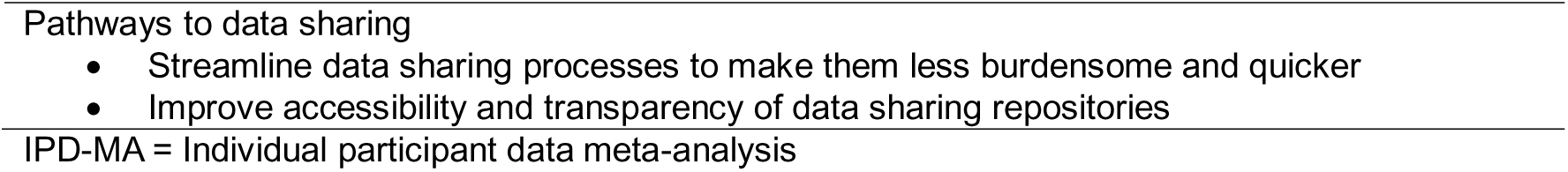
Consensus on key recommendations for authors, methodologists and educators of individual participant data meta-analyses of randomised trials

To maximise the advantages of IPD meta-analysis, the panel recommended more systematic identification and inclusion of unpublished evidence (routine trial registry searches and, where appropriate, seeking IPD for unpublished trials), greater use of IPD to assess data quality and trustworthiness, consideration of prospective meta-analysis approaches where feasible, and upfront evaluation of whether there is sufficient statistical power to justify an IPD project (particularly for interaction effects). The panel recommended appropriate analysis to account for missing data, appropriate adjustment for important prognostic factors, and, when IPD cannot be obtained for all eligible trials, explicit comparison of IPD and non-IPD evidence (using available aggregate data) including considering combining IPD and aggregate data where appropriate (e.g. in a sensitivity analysis).

For methodologists and educators, the recommendations are not simply to “summarise existing guidance”, but to make it easier to find, apply and enforce. The panel proposed an openly accessible IPD “toolbox” (building on www.ipdma.co.uk) that consolidates guidance into practical, usable resources – for example, templates for protocols and statistical analysis plans, worked examples of clearly reported one-stage and two-stage models (including interaction analyses that avoid aggregation bias), and checklists for data processing, missing data handling, and trustworthiness assessment. They also recommended targeted training and dissemination for authors, peer reviewers and journal editors, particularly in clinical and geographical areas where IPD meta-analysis is underused and using implementation science approaches to understand and overcome barriers to uptake of best practice. Finally, the panel highlighted priorities for future developments, including updating PRISMA-IPD to reflect current methodological expectations (e.g. reporting of trustworthiness checks and effect modification methods), and improving data-sharing pathways by streamlining processes and increasing the usability and transparency of repositories.

## Discussion

### Statement of principal findings

The number of IPD meta-analyses published annually has steadily increased over time but has plateaued in recent years. Most addressed questions in oncology and cardiovascular disease. The 100 recent pairwise IPD meta-analyses that we examined in detail were usually conducted in high-income countries, non-industry funded and non-Cochrane reviews. They tended to provide a clear rationale for their conduct, obtain data direct from trial investigators, include a high proportion of eligible trials and participants, assess risk of bias and cited reporting guidelines.

However, major shortcomings included a lack of transparency (i.e. limited protocols, statistical analysis plans and analysis code were available), notable flaws in conduct and analysis leading to potential bias and errors and major gaps in reporting. Additionally, opportunities were missed to search trial registers, include unpublished trials and utilise the IPD to its full advantage to inform or reduce risk of bias, conduct trustworthiness assessments, appropriately account for missing data, and separate out within-trial and across-trial information when assessing effect modification. Overall, these shortcomings suggest an underuse of existing methods and guidance.

The panel recommendations for IPD meta-analysis authors included increasing transparency through improved prospective registration, publishing full protocols and statistical analysis plans, clearly specifying statistical models and sharing analysis code. Authors are encouraged to improve reporting of their rationale for IPD meta-analysis and maximise searching for and, where appropriate, including unpublished evidence. Recommendations for IPD methodologists and educators include supporting improved implementation of existing guidance (such as through a new IPD toolbox), further developing methods in data processing and automation, and targeting capacity-building efforts toward geographical and clinical areas where IPD meta-analysis is currently underused.

### Strengths and limitations of the stud*y*

This study represents the most comprehensive search for IPD meta-analyses to date. We have provided an overview of the entire landscape of IPD meta-analysis of randomised trials and an in-depth review of the current state of play, spanning the spectrum of design, conduct, analysis and reporting. To our knowledge this is the first study to convene an international panel of IPD meta-analysis methodologists and experienced IPD meta-analysis authors to co-design the work and take part in a structured, consensus-based process to interpret the findings and identify priorities for improvement. This approach ensured that the recommendations were grounded in both our empirical results and current methodological expertise, and enabled us to develop actionable recommendations for the future of IPD meta-analysis, organised separately for authors, and for methodologists and educators.

A limitation of the in-depth review was that data were extracted in duplicate for only 40% of the IPD meta-analyses included. However, the impact of this on the results was likely minimal since we only moved from double to single extraction once agreement between reviewers was near-perfect. Our systematic review up to 2019 included only studies published in English, so we may have missed some IPD meta-analyses published in other languages. However, after 2019 we identified only two potentially eligible non-English full texts and neither were eligible, suggesting the number of IPD meta-analyses we may have missed is likely very small. Whilst we aimed for global representation among our consensus panel, there were no participants from low and middle-income countries. This reflects a wider issue of lack of IPD meta-analysis expertise in these regions.

### Comparison with other studies

Our findings are in line with Wang et al. (8), who found that only around one-third of IPD meta-analyses published until 2019 provided or mentioned a protocol. However, we found a substantial increase in pre-registration of IPD meta-analyses (e.g. on PROSPERO), yet, these registrations often provide insufficient detail on the design, conduct and analysis methods for an IPD meta-analysis. Whilst Simmonds et al. (5) found that few IPD meta-analyses published between 2008 and 2014 presented a risk of bias assessment or reported data checking, we found that the majority of recently published IPD meta-analyses did both, albeit with variations in reporting quality. However, we still found similar issues to Simmonds et al. (5) relating to poor and unclear reporting of statistical methods. For instance, the description of one-stage methods is often unclear to the point that multiple experienced extractors could not understand the approach. Compared to previous studies (9) (20), the separation of within- and across-trial information when undertaking participant-level subgroup analysis was improved in the set of recent IPD meta-analyses we reviewed, with 35/85 (41%) clearly separating this out and therefore not introducing aggregation bias. Although this represents progress, many IPD meta-analyses still fail to report this information clearly or continue to inappropriately conflate within- and across-trial information.

### Meaning of the study: possible explanations and implications for clinicians and policymakers, unanswered questions and future research

There appears to be a mismatch in our finding of high IPD retrieval rates and the panel’s recommendations to improve data sharing pathways. The panel raised overly complex and bureaucratic data sharing processes as one of the main barriers to conducting IPD meta-analysis (28). This apparent difference may have several explanations. First, by including only IPD meta-analyses that were successfully completed and published, we may have missed those abandoned due to difficulties obtaining sufficient IPD in a suitable timeframe (29), which is supported by examples shared by our panel and in published reports (30). Second, our data extraction relied on the self-reporting of IPD meta-analysis authors; which may not always be accurate. In some instances, IPD meta-analysis authors may introduce additional eligibility criteria (e.g. restricting inclusion to trials that shared IPD), thus artificially inflating data retrieval rates. Greater transparency, prospective registration and rigorous peer review may help address this in the future. Third, we did not collect data on the time taken to obtain IPD, which panel members noted takes considerable time and resource. Thus, improving data sharing pathways remains a priority. Data sharing repositories are far less utilised than anticipated, highlighting a clear need to improve their usability. In addition, data sharing agreements for investigator-to-investigator sharing need to be simplified and expedited.

A key consensus point was that IPD meta-analyses are largely conducted by a relatively small pool of researchers, mostly in high-income countries, and focus on a narrow range of clinical areas, despite global influence on guidelines, policy and clinical practice. Expanding capacity worldwide is essential to broadening the use and benefit of this methodology across the clinical spectrum. The panel propose an openly accessible online IPD toolbox as a simple solution to disseminate best-practice guidance to accelerate IPD meta-analysis methodology globally. However, this would require adequate funding, maintenance and regular updating to remain current and effective.

The consensus panel, which includes many long-standing Cochrane members, noted a low number of IPD meta-analyses published in the Cochrane Library and that these had decreased over time. This may reflect previously reported criticisms of Cochrane’s production tools and standards that lack the flexibility to accommodate IPD and prospective meta-analysis (31), and that many Cochrane review groups lack expertise in IPD. As Cochrane moves into a new era, with its 2025-2030 strategy (32) prioritising methodological innovation and promotion of health equity, engagement with the wider methods community alongside training and upskilling of Cochrane groups will be essential to allow re-integration of IPD meta-analysis methods for Cochrane reviews.

Medical research faces an integrity crisis, and trustworthiness issues in clinical trials often cannot be detected from publications alone (33). Access to IPD enables more rigorous checks of integrity, data quality and risk of bias, to ensure only the highest quality evidence informs policy and practice. Guidance and tools now exist on how to assess integrity in IPD meta-analysis (15), how to incorporate findings of IPD integrity assessment in synthesis (34) and how to use IPD to inform risk of bias assessments (35,36).

The panel found it encouraging that most IPD meta-analyses cited PRISMA-IPD (14), the main reporting guideline for IPD meta-analysis of randomised trials. However, major reporting gaps remain, likely reflecting methodological and reporting developments since PRISMA-IPD was published in 2015. Items such as whether and how trustworthiness of trials was assessed, whether and how within- and across-trial information were separated out in effect modification analyses, and whether there were prospective power calculations, are not included. This study provides a platform to guide an update to PRISMA-IPD. It is also important to encourage IPD meta-analyses to adopt PRISMA-IPD rather than other reporting guidelines (e.g. PRISMA 2020). This can be achieved through increasing training and awareness, as well as stricter enforcement by journal editors and peer reviewers. One such example includes the PRISMATIC project which is harmonising the PRISMA 2020 statement and several of its extensions (including PRISMA-IPD) and developing a web application which will allow authors to generate reporting guidance customised to the methods of their reviews (37).

## Conclusion

In the era of data sharing and personalised medicine, IPD meta-analyses are increasingly relevant for providing robust research findings: they can generate the highest quality evidence to inform clinical guidelines, policy and ultimately patient care. Through our review and consensus process, we have identified areas for improvement, including the need for greater transparency; improved conduct, analysis and reporting; capacity building and wider education; and better pathways to data sharing. Implementing these recommendations will help realise the full potential of IPD meta-analysis, to generate robust evidence to inform clinical decision making worldwide.

## Footnotes

### Data and code sharing

All data and statistical analysis code underlying this study is publicly available on the Open Science Framework (1), or within the supplementary material.

### Ethics approval

Ethics approval was not required for this study as all data in parts 1 and 2 were already in the public domain, and results from part 3 were simply interpretation and discussion points by authors or acknowledged contributors.

### Transparency statement

The lead authors (ALS and PJG) affirm that the manuscript is an honest, accurate, and transparent account of the study being reported; that no important aspects of the study have been omitted; and that any discrepancies from the study as originally planned and registered have been explained.

### Contributor statement

ALS and PJG conceived the study. ALS, JA, LSN, KEH, RB, DGH, TL, NM, DN, RDR, LHMR, MS, LAS, WT, JFT, RW and PJG developed the study protocol. KEH carried out the searches. ALS, JA, LSN, KEH, TL, DN, SL, JS and PJG screened studies. ALS, JA, KEH, LHMR and PJG developed and piloted the data extraction tool. ALS, JA, LSN, KEH, RB, DGH, TL, NM, DN, LHMR, WT, RW and PJG extracted data. ALS, JA, LSN, KEH and PJG carried out the analysis. ALS, JA and PJG designed the consensus workshops. KEH, DGH, NM, RDR, LHMR, MS, LAS, WT, JFT, RW, AA, MB, SB, JE, MH, SL, YL, SS, LKS, KIES, JS, CLV, IRW and JGW actively participated in the consensus workshops, JA, LSN and DN took detailed minutes, ALS and PJG chaired the workshop groups. RB could not take part in the consensus workshop but provided detailed written comments. ALS and PJG drafted the manuscript. All authors critically revised the manuscript across several iterations for important intellectual content and gave final approval for the article.

ALS and PJG are the guarantors and accept full responsibility for the work and the conduct of the study, had access to the data, and controlled the decision to publish. The corresponding authors attest that all listed authors meet authorship criteria and that no others meeting the criteria have been omitted.

### Competing interests declaration

All authors have completed the ICMJE uniform disclosure form at http://www.icmje.org/disclosure-of-interest/ and declare: RB is chair of the data monitoring committee for the Predict & Prevent AECOPD Trial and a member of the Arthritis UK College of Experts, RDR is the creator of www.ipd.co.uk, RDR, JFT and LAS are editors of Individual Participant Data Meta-Analysis: A Handbook for Healthcare Research, for which they receive royalties for book sales; no other relationships or activities that could appear to have influenced the submitted work.

## Funding

This research was supported by a University of Sydney – University College London Ignition Grant. The funders had no role in the study design; in the collection, analysis, and interpretation of data; in the writing of the report; and in the decision to submit the article for publication.

LSN is supported by a National Institute for Health and Care Research (NIHR) Doctoral Fellowship (NIHR306441). LHMR, JFT, SB, CLV, IRW and PJG are supported by the UK Medical Research Council (MC_UU_00004/06). PJG is also supported by a Prostate Cancer Foundation-John Black Charitable Foundation Young Investigator Award. RDR is a NIHR Senior Investigator. RDR, MH, JE and KIES are supported by an MRC-NIHR grant (MR/V038168/1) and the NIHR Birmingham Biomedical Research Centre at the University Hospitals Birmingham NHS Foundation Trust and the University of Birmingham. LAS is the Director of the NIHR Evidence Synthesis Programme for which her institution receives an honorarium payment. The views expressed are those of the authors and not necessarily those of the NHS, the NIHR or the Department of Health and Social Care. AA is supported by a Swiss National Science Foundation postdoc mobility grant (#P500PM_221961).

## Data Availability

https://osf.io/zw6mf

## Acknowledgements

We thank Lilly Sophie Andreas (University Medical Centre Rostock) for technical support, Nipun Shrestha, Yuetong Ren and Talia Palacios (all University of Sydney) for assistance with screening records.

