## Supplementary Material for "State of play in individual participant data meta-analyses of randomised trials: Systematic review and consensus-based recommendations"

### Supplementary File 1: PRISMA 2020 checklist

| **Section and Topic** | **Item #** | **Checklist item** | **Location where item is reported** |
| --- | --- | --- | --- |
| **TITLE** | | |  |
| Title | 1 | Identify the report as a systematic review. | Page 1 |
| **ABSTRACT** | | |  |
| Abstract | 2 | See the PRISMA 2020 for Abstracts checklist. |  |
| **INTRODUCTION** | | |  |
| Rationale | 3 | Describe the rationale for the review in the context of existing knowledge. | Page 5 |
| Objectives | 4 | Provide an explicit statement of the objective(s) or question(s) the review addresses. | Page 5 |
| **METHODS** | | |  |
| Eligibility criteria | 5 | Specify the inclusion and exclusion criteria for the review and how studies were grouped for the syntheses. | Page 6 |
| Information sources | 6 | Specify all databases, registers, websites, organisations, reference lists and other sources searched or consulted to identify studies. Specify the date when each source was last searched or consulted. | Page 6, Supplementary File 3 |
| Search strategy | 7 | Present the full search strategies for all databases, registers and websites, including any filters and limits used. | Page 6, Supplementary File 3 |
| Selection process | 8 | Specify the methods used to decide whether a study met the inclusion criteria of the review, including how many reviewers screened each record and each report retrieved, whether they worked independently, and if applicable, details of automation tools used in the process. | Page 6 |
| Data collection process | 9 | Specify the methods used to collect data from reports, including how many reviewers collected data from each report, whether they worked independently, any processes for obtaining or confirming data from study investigators, and if applicable, details of automation tools used in the process. | Pages 6-7 |
| Data items | 10a | List and define all outcomes for which data were sought. Specify whether all results that were compatible with each outcome domain in each study were sought (e.g. for all measures, time points, analyses), and if not, the methods used to decide which results to collect. | Pages 6-7, Supplementary Files 4 & 5 |
|  | 10b | List and define all other variables for which data were sought (e.g. participant and intervention characteristics, funding sources). Describe any assumptions made about any missing or unclear information. | Pages 6-7, Supplementary Files 4 & 5 |
| Study risk of bias assessment | 11 | Specify the methods used to assess risk of bias in the included studies, including details of the tool(s) used, how many reviewers assessed each study and whether they worked independently, and if applicable, details of automation tools used in the process. | NA |
| Effect measures | 12 | Specify for each outcome the effect measure(s) (e.g. risk ratio, mean difference) used in the synthesis or presentation of results. | NA |
| Synthesis methods | 13a | Describe the processes used to decide which studies were eligible for each synthesis (e.g. tabulating the study intervention characteristics and comparing against the planned groups for each synthesis (item #5)). | NA |
|  | 13b | Describe any methods required to prepare the data for presentation or synthesis, such as handling of missing summary statistics, or data conversions. | NA |
|  | 13c | Describe any methods used to tabulate or visually display results of individual studies and syntheses. | NA |
|  | 13d | Describe any methods used to synthesize results and provide a rationale for the choice(s). If meta-analysis was performed, describe the model(s), method(s) to identify the presence and extent of statistical heterogeneity, and software package(s) used. | NA |
|  | 13e | Describe any methods used to explore possible causes of heterogeneity among study results (e.g. subgroup analysis, meta-regression). | NA |
|  | 13f | Describe any sensitivity analyses conducted to assess robustness of the synthesized results. | NA |
| Reporting bias assessment | 14 | Describe any methods used to assess risk of bias due to missing results in a synthesis (arising from reporting biases). | NA |
| Certainty assessment | 15 | Describe any methods used to assess certainty (or confidence) in the body of evidence for an outcome. | NA |
| **RESULTS** | | |  |
| Study selection | 16a | Describe the results of the search and selection process, from the number of records identified in the search to the number of studies included in the review, ideally using a flow diagram. | Page 9, Supplementary Figure 1 |
|  | 16b | Cite studies that might appear to meet the inclusion criteria, but which were excluded, and explain why they were excluded. | Supplementary Figure 1 |
| Study characteristics | 17 | Cite each included study and present its characteristics. | NA |
| Risk of bias in studies | 18 | Present assessments of risk of bias for each included study. | NA |
| Results of individual studies | 19 | For all outcomes, present, for each study: (a) summary statistics for each group (where appropriate) and (b) an effect estimate and its precision (e.g. confidence/credible interval), ideally using structured tables or plots. | NA |
| Results of syntheses | 20a | For each synthesis, briefly summarise the characteristics and risk of bias among contributing studies. | NA |
|  | 20b | Present results of all statistical syntheses conducted. If meta-analysis was done, present for each the summary estimate and its precision (e.g. confidence/credible interval) and measures of statistical heterogeneity. If comparing groups, describe the direction of the effect. | NA |
|  | 20c | Present results of all investigations of possible causes of heterogeneity among study results. | NA |
|  | 20d | Present results of all sensitivity analyses conducted to assess the robustness of the synthesized results. | NA |
| Reporting biases | 21 | Present assessments of risk of bias due to missing results (arising from reporting biases) for each synthesis assessed. | NA |
| Certainty of evidence | 22 | Present assessments of certainty (or confidence) in the body of evidence for each outcome assessed. | NA |
| **DISCUSSION** | | |  |
| Discussion | 23a | Provide a general interpretation of the results in the context of other evidence. | Page 15 |
|  | 23b | Discuss any limitations of the evidence included in the review. | Pages 14-15 |
|  | 23c | Discuss any limitations of the review processes used. | Pages 14-15 |
|  | 23d | Discuss implications of the results for practice, policy, and future research. | Pages 15-17 |
| **OTHER INFORMATION** | | |  |
| Registration and protocol | 24a | Provide registration information for the review, including register name and registration number, or state that the review was not registered. | Page 6 |
|  | 24b | Indicate where the review protocol can be accessed, or state that a protocol was not prepared. | Page 6, and references |
|  | 24c | Describe and explain any amendments to information provided at registration or in the protocol. | Page 7 |
| Support | 25 | Describe sources of financial or non-financial support for the review, and the role of the funders or sponsors in the review. | Page 19 |
| Competing interests | 26 | Declare any competing interests of review authors. | Pages 18-19 |
| Availability of data, code and other materials | 27 | Report which of the following are publicly available and where they can be found: template data collection forms; data extracted from included studies; data used for all analyses; analytic code; any other materials used in the review. | Page 18 |

### Supplementary File 2: ACCORD checklist

| Item  No. | Section | Checklist Item (*help text*) | Page  No. |
| --- | --- | --- | --- |
| T1 | **Title** | Identify the article as reporting a consensus exercise and state the consensus methods used in the title.  *For example, Delphi or nominal group technique.* | 1 |
| I1 | **Introduction** | Explain why a consensus exercise was chosen over other approaches. | - |
| I2 |  | State the aim of the consensus exercise, including its intended audience and geographical scope (national, regional, global). | 5 |
| I3 |  | If the consensus exercise is an update of an existing document, state why an update is needed, and provide the citation for the original document. | NA |
| M1 | **Methods**  Registration | If the study or study protocol was prospectively registered, state the registration platform and provide a link. If the exercise was not registered, this should be stated.  *Recommended to include the date of registration.* | 6 |
| M2 | Selection of SC and/or panellists | Describe the role(s) and areas of expertise or experience of those directing the consensus exercise.  *For example, whether the project was led by a chair, co-chairs or a steering committee, and, if so, how they were chosen. List their names if appropriate, and whether there were any subgroups for individual steps in the process.* | 7, 9 |
| M3 |  | Explain the criteria for panellist inclusion and the rationale for panellist numbers. State who was responsible for panellist selection. | 8 |
| M4 |  | Describe the recruitment process (how panellists were invited to participate).  *Include communication/advertisement method(s) and locations, numbers of invitations sent, and whether there was centralised oversight of invitations or if panellists were asked/allowed to suggest other members of the panel.* | 8 |
| M5 |  | Describe the role of any members of the public, patients or carers in the different steps of the study. | 9 |
| M6 | Preparatory research | Describe how information was obtained prior to generating items or other materials used during the consensus exercise.  *This might include a literature review, interviews, surveys, or another process.* | 5-7 |
| M7 |  | Describe any systematic literature search in detail, including the search strategy and dates of search or the citation if published already.  *Provide the details suggested by the reporting guideline PRISMA and the related PRISMA-Search extension.* | 6-7 |
| M8 |  | Describe how any existing scientific evidence was summarised and if this evidence was provided to the panellists. | Supp File 6 |
| M9 | Assessing consensus | Describe the methods used and steps taken to gather panellist input and reach consensus (for example, Delphi, RAND-UCLA, nominal group technique).  *If modifications were made to the method in its original form, provide a detailed explanation of how the method was adjusted and why this was necessary for the purpose of your consensus-based study.* | 8-9 |
| M10 |  | Describe how each question or statement was presented and the response options. State whether panellists were able to or required to explain their responses, and whether they could propose new items.  *Where possible, present the questionnaire or list of statements as supplementary material.* | Supp File 6 |
| M11 |  | State the objective of each consensus step.  *A step could be a consensus meeting, a discussion or interview session, or a Delphi round.* | 8, Supp File 6 |
| M12 |  | State the definition of consensus (for example, number, percentage, or categorical rating, such as ‘agree’ or ‘strongly agree’) and explain the rationale for that definition. | 8-9 |
| M13 |  | State whether items that met the prespecified definition of consensus were included in any subsequent voting rounds. | NA |
| M14 |  | For each step, describe how responses were collected, and whether responses were collected in a group setting or individually. | 8 |
| M15 |  | Describe how responses were processed and/or synthesised.  *Include qualitative analyses of free-text responses (for example, thematic, content or cluster analysis) and/or quantitative analytical methods, if used.* | 9 |
| M16 |  | Describe any piloting of the study materials and/or survey instruments.  *Include how many individuals piloted the study materials, the rationale for the selection of those individuals, any changes made as a result and whether their responses were used in the calculation of the final consensus. If no pilot was conducted, this should be stated.* | 9 |
| M17 |  | If applicable, describe how feedback was provided to panellists at the end of each consensus step or meeting.  *State whether feedback was quantitative (for example, approval rates per topic/item) and/or qualitative (for example, comments, or lists of approved items), and whether it was anonymised.* | NA |
| M18 |  | State whether anonymity was planned in the study design. Explain where and to whom it was applied and what methods were used to guarantee anonymity. | - |
| M19 |  | State if the steering committee was involved in the decisions made by the consensus panel.  *For example, whether the steering committee or those managing consensus also had voting rights.* | - |
| M20 | Participation | Describe any incentives used to encourage responses or participation in the consensus process.  *For example, were invitations to participate reiterated, or were participants reimbursed for their time.* | 12 |
| M21 |  | Describe any adaptations to make the surveys/meetings more accessible.  *For example, the languages in which the surveys/meetings were conducted and whether translations or plain language summaries were available*. | NA |
| R1 | Results | State when the consensus exercise was conducted. List the date of initiation and the time taken to complete each consensus step, analysis, and any extensions or delays in the analysis. | 11 |
| R2 |  | Explain any deviations from the study protocol, and why these were necessary.  *For example, addition of panel members during the exercise, number of consensus steps, stopping criteria; report the step(s) in which this occurred.* | NA |
| R3 |  | For each step, report quantitative (number of panellists, response rate) and qualitative (relevant socio-demographics) data to describe the participating panellists. | 12, Supp Table 7 |
| R4 |  | Report the final outcome of the consensus process as qualitative (for example, aggregated themes from comments) and/or quantitative (for example, summary statistics, score means, medians and/or ranges) data. | Table 2, Table 3 |
| R5 |  | List any items or topics that were modified or removed during the consensus process. Include why and when in the process they were modified or removed. | NA |
| D1 | Discussion | Discuss the methodological strengths and limitations of the consensus exercise.  *Include factors that may have impacted the decisions (for example, response rates, representativeness of the panel, potential for feedback during consensus to bias responses, potential impact of any non-anonymised interactions).* | 14-15 |
| D2 |  | Discuss whether the recommendations are consistent with any pre-existing literature and, if not, propose reasons why this process may have arrived at alternative conclusions. | 15-17 |
| O1 | Other information | List any endorsing organisations involved and their role. | NA |
| O2 |  | State any potential conflicts of interests, including among those directing the consensus study and panellists. Describe how conflicts of interest were managed. | 18-19 |
| O3 |  | State any funding received and the role of the funder.  *Specify, for example, any funder involvement in the study concept/design, participation in the steering committee, conducting the consensus process, funding of any medical writing support. This could be disclosed in the methods or in the relevant transparency section of the manuscript. Where a funder did not play a role in the process or influence the decisions reached, this should be specified.* | 19 |

### Supplementary File 3: Search strategy

#### Search results (overall)

| **Database searched** | **Records retrieved** |
| --- | --- |
| Search 1 (06/07/2022) | 2831 |
| Search 2 (26/02/2024) | 2173 |
| Total | 5004 |
| Duplicates detected in Covidence | 416 |
| **Total after deduplication in Covidence** | **4588** |

#### Search 1 – July 2022

Date of search: 06/07/2022

| **Database searched** | **Records retrieved** |
| --- | --- |
| Ovid MEDLINE | 1,278 |
| Embase | 2,610 |
| EBM Reviews | 357 |
| Total | 4245 |
| Duplicates removed in Endnote | 1414 |
| **Total after deduplication in Endnote** | **2831** |

**Database:**
Ovid MEDLINE(R) ALL <1946 to July 05, 2022>

| **#** | **Query** | **Results** |
| --- | --- | --- |
| 1 | (individual patient$ adj6 data).ti,ab. | 5,595 |
| 2 | (individual patient$ adj6 report$).ti,ab. | 533 |
| 3 | (individual patient$ adj6 outcome$).ti,ab. | 1,615 |
| 4 | (individual patient$ adj6 level$).ti,ab. | 2,324 |
| 5 | individual participant data.ti,ab. | 1,530 |
| 6 | (individual subject$ adj6 data).ti,ab. | 430 |
| 7 | (individual subject$ adj6 report$).ti,ab. | 45 |
| 8 | (individual subject$ adj6 outcome$).ti,ab. | 35 |
| 9 | (individual subject$ adj6 level$).ti,ab. | 372 |
| 10 | (raw patient$ adj6 data).ti,ab. | 18 |
| 11 | ipd.ti,ab. | 4,336 |
| 12 | (raw patient$ adj6 level$).ti,ab. | 2 |
| 13 | (raw subject$ adj6 data).ti,ab. | 2 |
| 14 | idiopathic.ti,ab. | 132,829 |
| 15 | immediate pigment darkening.ti,ab. | 52 |
| 16 | intermittent peritoneal dialysis.ti,ab. | 414 |
| 17 | invasive pneumococcal disease.ti,ab. | 2,671 |
| 18 | indirect photometric detection.ti,ab. | 77 |
| 19 | interaural phase disparity.ti,ab. | 11 |
| 20 | 1 or 2 or 3 or 4 or 5 or 6 or 7 or 8 or 9 or 10 or 11 or 12 or 13 | 14,792 |
| 21 | 14 or 15 or 16 or 17 or 18 or 19 | 136,049 |
| 22 | 20 not 21 | 12,585 |
| 23 | meta analysis.mp,pt. | 247,924 |
| 24 | search:.tw. | 578,651 |
| 25 | review.pt. | 3,009,159 |
| 26 | 23 or 24 or 25 | 3,451,418 |
| 27 | 22 and 26 | 5,621 |
| 28 | limit 27 to humans | 4,949 |
| 29 | limit 28 to dt=20190801-20220706 | 1,278 |

**Database:**
Embase Classic+Embase <1947 to 2022 July 05>

| **#** | **Query** | **Results** |
| --- | --- | --- |
| 1 | (individual patient$ adj6 data).ti,ab. | 9,218 |
| 2 | (individual patient$ adj6 report$).ti,ab. | 871 |
| 3 | (individual patient$ adj6 outcome$).ti,ab. | 2,425 |
| 4 | (individual patient$ adj6 level$).ti,ab. | 3,802 |
| 5 | individual participant data.ti,ab. | 1,984 |
| 6 | (individual subject$ adj6 data).ti,ab. | 585 |
| 7 | (individual subject$ adj6 report$).ti,ab. | 68 |
| 8 | (individual subject$ adj6 outcome$).ti,ab. | 51 |
| 9 | (individual subject$ adj6 level$).ti,ab. | 498 |
| 10 | (raw patient$ adj6 data).ti,ab. | 28 |
| 11 | ipd.ti,ab. | 7,014 |
| 12 | (raw patient$ adj6 level$).ti,ab. | 2 |
| 13 | (raw subject$ adj6 data).ti,ab. | 2 |
| 14 | idiopathic.ti,ab. | 204,388 |
| 15 | immediate pigment darkening.ti,ab. | 72 |
| 16 | intermittent peritoneal dialysis.ti,ab. | 540 |
| 17 | invasive pneumococcal disease.ti,ab. | 3,444 |
| 18 | indirect photometric detection.ti,ab. | 152 |
| 19 | interaural phase disparity.ti,ab. | 11 |
| 20 | 1 or 2 or 3 or 4 or 5 or 6 or 7 or 8 or 9 or 10 or 11 or 12 or 13 | 23,136 |
| 21 | 14 or 15 or 16 or 17 or 18 or 19 | 208,601 |
| 22 | 20 not 21 | 19,903 |
| 23 | meta analysis.mp,pt. | 364,927 |
| 24 | search:.tw. | 734,332 |
| 25 | review.pt. | 2,968,726 |
| 26 | 23 or 24 or 25 | 3,627,310 |
| 27 | 22 and 26 | 7,540 |
| 28 | limit 27 to humans | 7,137 |
| 29 | limit 28 to yr="2019 -Current" | 2,610 |

**Database:**
EBM Reviews - Cochrane Database of Systematic Reviews <2005 to June 29, 2022>
EBM Reviews - ACP Journal Club <1991 to June 2022>
EBM Reviews - Database of Abstracts of Reviews of Effects <1st Quarter 2016>
EBM Reviews - Cochrane Clinical Answers <June 2022>
EBM Reviews - Cochrane Central Register of Controlled Trials <June 2022>
EBM Reviews - Cochrane Methodology Register <3rd Quarter 2012>
EBM Reviews - Health Technology Assessment <4th Quarter 2016>
EBM Reviews - NHS Economic Evaluation Database <1st Quarter 2016>

| **#** | **Query** | **Results** |
| --- | --- | --- |
| 1 | (individual patient$ adj6 data).ti,ab. | 2,029 |
| 2 | (individual patient$ adj6 report$).ti,ab. | 95 |
| 3 | (individual patient$ adj6 outcome$).ti,ab. | 278 |
| 4 | (individual patient$ adj6 level$).ti,ab. | 497 |
| 5 | individual participant data.ti,ab. | 246 |
| 6 | (individual subject$ adj6 data).ti,ab. | 65 |
| 7 | (individual subject$ adj6 report$).ti,ab. | 6 |
| 8 | (individual subject$ adj6 outcome$).ti,ab. | 15 |
| 9 | (individual subject$ adj6 level$).ti,ab. | 60 |
| 10 | (raw patient$ adj6 data).ti,ab. | 1 |
| 11 | ipd.ti,ab. | 677 |
| 12 | (raw patient$ adj6 level$).ti,ab. | 0 |
| 13 | (raw subject$ adj6 data).ti,ab. | 1 |
| 14 | idiopathic.ti,ab. | 11,620 |
| 15 | immediate pigment darkening.ti,ab. | 11 |
| 16 | intermittent peritoneal dialysis.ti,ab. | 29 |
| 17 | invasive pneumococcal disease.ti,ab. | 177 |
| 18 | indirect photometric detection.ti,ab. | 0 |
| 19 | interaural phase disparity.ti,ab. | 0 |
| 20 | 1 or 2 or 3 or 4 or 5 or 6 or 7 or 8 or 9 or 10 or 11 or 12 or 13 | 3,261 |
| 21 | 14 or 15 or 16 or 17 or 18 or 19 | 11,837 |
| 22 | 20 not 21 | 3,112 |
| 23 | meta analysis.mp,pt. | 55,125 |
| 24 | search:.tw. | 57,504 |
| 25 | review.pt. | 3,122 |
| 26 | 23 or 24 or 25 | 88,461 |
| 27 | 22 and 26 | 1,311 |
| 28 | limit 27 to humans [Limit not valid in CDSR,ACP Journal Club,DARE,CCA,CCTR,CLCMR; records were retained] | 1,306 |
| 29 | limit 28 to yr="2019 -Current" [Limit not valid in DARE; records were retained] | 357 |

#### Search 2 – February 2024

**Date of search:** 26/02/2024

| **Database searched** | **Records retrieved** |
| --- | --- |
| Ovid MEDLINE | 764 |
| Embase | 1886 |
| EBM Reviews | 280 |
| Total | 2930 |
| Duplicates removed in Endnote | 757 |
| **Total after deduplication in Endnote** | **2173** |

**Database:** 
Ovid MEDLINE(R) ALL <1946 to February 23, 2024>

| **#** | **Query** | **Results** |
| --- | --- | --- |
| 1 | (individual patient$ adj6 data).ti,ab. | 6462 |
| 2 | (individual patient$ adj6 report$).ti,ab. | 598 |
| 3 | (individual patient$ adj6 outcome$).ti,ab. | 1809 |
| 4 | (individual patient$ adj6 level$).ti,ab. | 2629 |
| 5 | individual participant data.ti,ab. | 1985 |
| 6 | (individual subject$ adj6 data).ti,ab. | 441 |
| 7 | (individual subject$ adj6 report$).ti,ab. | 46 |
| 8 | (individual subject$ adj6 outcome$).ti,ab. | 38 |
| 9 | (individual subject$ adj6 level$).ti,ab. | 399 |
| 10 | (raw patient$ adj6 data).ti,ab. | 24 |
| 11 | ipd.ti,ab. | 4983 |
| 12 | (raw patient$ adj6 level$).ti,ab. | 3 |
| 13 | (raw subject$ adj6 data).ti,ab. | 2 |
| 14 | idiopathic.ti,ab. | 143528 |
| 15 | immediate pigment darkening.ti,ab. | 53 |
| 16 | intermittent peritoneal dialysis.ti,ab. | 418 |
| 17 | invasive pneumococcal disease.ti,ab. | 2880 |
| 18 | indirect photometric detection.ti,ab. | 78 |
| 19 | interaural phase disparity.ti,ab. | 11 |
| 20 | 1 or 2 or 3 or 4 or 5 or 6 or 7 or 8 or 9 or 10 or 11 or 12 or 13 | 16913 |
| 21 | 14 or 15 or 16 or 17 or 18 or 19 | 146963 |
| 22 | 20 not 21 | 14497 |
| 23 | meta analysis.mp,pt. | 298668 |
| 24 | search:.tw. | 682659 |
| 25 | review.pt. | 3282844 |
| 26 | 23 or 24 or 25 | 3794553 |
| 27 | 22 and 26 | 6597 |
| 28 | limit 27 to humans | 5772 |
| 29 | limit 28 to dt=20220601-20240226 | 764 |

**Database:** 
Embase Classic+Embase <1947 to 2024 February 23>

| **#** | **Query** | **Results** |
| --- | --- | --- |
| 1 | (individual patient$ adj6 data).ti,ab. | 10840 |
| 2 | (individual patient$ adj6 report$).ti,ab. | 991 |
| 3 | (individual patient$ adj6 outcome$).ti,ab. | 2771 |
| 4 | (individual patient$ adj6 level$).ti,ab. | 4430 |
| 5 | individual participant data.ti,ab. | 2627 |
| 6 | (individual subject$ adj6 data).ti,ab. | 616 |
| 7 | (individual subject$ adj6 report$).ti,ab. | 70 |
| 8 | (individual subject$ adj6 outcome$).ti,ab. | 55 |
| 9 | (individual subject$ adj6 level$).ti,ab. | 541 |
| 10 | (raw patient$ adj6 data).ti,ab. | 35 |
| 11 | ipd.ti,ab. | 8315 |
| 12 | (raw patient$ adj6 level$).ti,ab. | 3 |
| 13 | (raw subject$ adj6 data).ti,ab. | 2 |
| 14 | idiopathic.ti,ab. | 223282 |
| 15 | immediate pigment darkening.ti,ab. | 73 |
| 16 | intermittent peritoneal dialysis.ti,ab. | 558 |
| 17 | invasive pneumococcal disease.ti,ab. | 3736 |
| 18 | indirect photometric detection.ti,ab. | 153 |
| 19 | interaural phase disparity.ti,ab. | 11 |
| 20 | 1 or 2 or 3 or 4 or 5 or 6 or 7 or 8 or 9 or 10 or 11 or 12 or 13 | 26977 |
| 21 | 14 or 15 or 16 or 17 or 18 or 19 | 227807 |
| 22 | 20 not 21 | 23397 |
| 23 | meta analysis.mp,pt. | 437738 |
| 24 | search:.tw. | 861445 |
| 25 | review.pt. | 3233861 |
| 26 | 23 or 24 or 25 | 3991620 |
| 27 | 22 and 26 | 8987 |
| 28 | limit 27 to humans | 8561 |
| 29 | limit 28 to yr="2022 -Current" | 1886 |

**Databases (EBM – All):** 
EBM Reviews - Cochrane Database of Systematic Reviews <2005 to February 21, 2024> 
EBM Reviews - ACP Journal Club <1991 to February 2024> 
EBM Reviews - Database of Abstracts of Reviews of Effects <1st Quarter 2016> 
EBM Reviews - Cochrane Clinical Answers <February 2024> 
EBM Reviews - Cochrane Central Register of Controlled Trials <January 2024> 
EBM Reviews - Cochrane Methodology Register <3rd Quarter 2012> 
EBM Reviews - Health Technology Assessment <4th Quarter 2016> 
EBM Reviews - NHS Economic Evaluation Database <1st Quarter 2016>

| **#** | **Query** | **Results** |
| --- | --- | --- |
| 1 | (individual patient$ adj6 data).ti,ab. | 2055 |
| 2 | (individual patient$ adj6 report$).ti,ab. | 96 |
| 3 | (individual patient$ adj6 outcome$).ti,ab. | 269 |
| 4 | (individual patient$ adj6 level$).ti,ab. | 542 |
| 5 | individual participant data.ti,ab. | 273 |
| 6 | (individual subject$ adj6 data).ti,ab. | 61 |
| 7 | (individual subject$ adj6 report$).ti,ab. | 9 |
| 8 | (individual subject$ adj6 outcome$).ti,ab. | 15 |
| 9 | (individual subject$ adj6 level$).ti,ab. | 63 |
| 10 | (raw patient$ adj6 data).ti,ab. | 3 |
| 11 | ipd.ti,ab. | 695 |
| 12 | (raw patient$ adj6 level$).ti,ab. | 0 |
| 13 | (raw subject$ adj6 data).ti,ab. | 1 |
| 14 | idiopathic.ti,ab. | 12224 |
| 15 | immediate pigment darkening.ti,ab. | 11 |
| 16 | intermittent peritoneal dialysis.ti,ab. | 30 |
| 17 | invasive pneumococcal disease.ti,ab. | 170 |
| 18 | indirect photometric detection.ti,ab. | 0 |
| 19 | interaural phase disparity.ti,ab. | 0 |
| 20 | 1 or 2 or 3 or 4 or 5 or 6 or 7 or 8 or 9 or 10 or 11 or 12 or 13 | 3378 |
| 21 | 14 or 15 or 16 or 17 or 18 or 19 | 12435 |
| 22 | 20 not 21 | 3237 |
| 23 | meta analysis.mp,pt. | 50861 |
| 24 | search:.tw. | 49959 |
| 25 | review.pt. | 0 |
| 26 | 23 or 24 or 25 | 77727 |
| 27 | 22 and 26 | 1257 |
| 28 | limit 27 to humans [Limit not valid in CDSR,ACP Journal Club,DARE,CCA,CCTR,CLCMR; records were retained] | 1252 |
| 29 | limit 28 to yr="2022 -Current" [Limit not valid in DARE; records were retained] | 280 |

### Supplementary File 4: Variables extracted in Part 1: Systematic review

For analysis of IPD-MAs over time, we included the full sample of identified IPD-MAs, and extracted the following variables:

| **Variable** | **Definition** |
| --- | --- |
| Year of publication | Where first published if multiple outlets |
| Subject area | Categories from the International Classification of Diseases 11^th^ Revision |
| Type of journal | Q1, Q2, Q3, Q4, no impact factor.  Based on the Journal Citation Reports (JCR, based on 2023 impact factor) |
| Geographical location of corresponding author | Determined by affiliation or other information available in publication |
| Number of trials for which IPD was obtained | As stated in publication or supplementary material |
| Number of participants for which IPD was obtained | As stated in publication or supplementary material |
| Registration of IPD-MA | yes - PROSPERO, yes – elsewhere, not stated |
| Analysis type | Pairwise, network, both |
| Update of previous study? | Yes, no |

### Supplementary File 5: Extraction guide for Part 2: In-depth review

| Field | Guidance |
| --- | --- |
| General  characteristics |  |
| Named collaboration?  If 'Yes' override w. text | Control + F for collaboration and consortium  Quick review of abstract and methods for collaboration name  Also check author list – to see if it is published on behalf of a collaboration |
| Cochrane  review? | Published with Cochrane? (yes/no) |
| Source of  funding | Note this should be the source of funding for the meta-analysis, not the source of funding for the individual studies.  Copy and paste funding text |
| Searching  trial registries | Common trial registries include:   - ClinicalTrials.gov - ISRCTN - ANZCTR - EU-CTR - WHO ICTRP   For a full list of data providers to the WHO ICTRP, see <https://www.who.int/clinical-trials-registry-platform/network/data-providers>  Note that searches of CENTRAL without also listing a trial registry listed should be coded as No. |
| Prospective/  retrospective meta-analysis | Note that prospectively registered does not necessarily mean it is a prospective meta-analysis (often it is retrospective still).  Definition of a prospective meta-analysis:  The key feature of a prospective meta-analysis (PMA) is that the studies or cohorts are identified as eligible for inclusion in the meta-analysis, and hypotheses and analysis strategies are specified, before the results of the studies or cohorts related to the PMA research question are known. (<https://www.bmj.com/content/367/bmj.l5342> )  Check if studies were identified before their results are known. If not stated, usually retrospective.  Definition of nested PMA: “A nested PMA integrates prospective evidence into a retrospective meta-analysis, making best use of existing and emerging evidence while also retaining some benefits of PMAs.” (Seidler et al. BMJ 2019) |
| Data  retrieval |  |
| Number of eligible  identified trials | Look at flow diagram (usually Fig 1)  All trials that were eligible according to PICO criteria, i.e. if a trial was deemed not eligible since it did not share IPD, it should still be counted here.  If a trial was later excluded due to integrity or data quality issues it should still be counted here.  Only count trials that are completed at time of analysis (not trials that could not be included since they were still ongoing) – this is particularly relevant for prospective meta-analyses. |
| Number of eligible  identified participants | If this is not provided in the paper, note this as ‘not stated’.  Eligible as defined in the meta-analysis (not necessarily the trial) |
| Number of eligible trials for which IPD were obtained | This is data that was extracted from Part 1 – check this is correct. |
| Number of eligible participants  for which IPD were obtained | This is data that was extracted from Part 1 – check this is correct. |
| Data obtained from: | If investigators were contacted, but it is not directly stated that they were the ones providing data, it can be assumed data were obtained from investigators, unless it is directly stated that data is obtained from other sources (e.g., repository).  If data are accessed remotely, we assume that it is from a data repository. |
| If data was obtained from  repositories, was it downloadable? | Downloadable meaning: Could the IPD be downloaded and accessed on a local drive and be merged with other IPD datasets that were received directly from investigators.  Some repositories only let you access the data within the platform and download aggregate-level data/results. This means that you cannot do one-stage analyses and would be classified as ‘not downloadable’. |
| Number of trials with AD extracted | This refers to trials for which **no IPD were available** and only AD were extracted  Please write “0” if no trials with AD were extracted |
| Number of participants with AD extracted | This refers to trials for which **no IPD were available** and only AD were extracted  Please write “0” if no participants with AD were extracted |
| Were unpublished  trials obtained? | Did they include unpublished trials (not did they intend to in case they identified some). This field is trying to capture whether the IPD meta-analysis managed to include trials that were completed but not published in a journal, and therefore help to mitigate publication bias. |
| Unpublished trials number  (How many) | If not clearly stated, mark this as ‘unclear’ |
| Unpublished trial participants  (How many) | If not clearly stated, mark this as ‘unclear’ |
| Reporting and conduct  of IPD-MAs |  |
| Rationale for conducting an IPD-MA | This will frequently be reported in the introduction section, but sometimes in the methods or discussion section. Note that this should be specific to their study, not just general text like “IPD-MA are good for X, Y, Z reason”.  We are looking for a clear rationale and not a general statement. If needed, copy the free-text and we can review centrally. |
| IPD rationale  (choose all that apply) | This may be challenging and may at times need a second opinion. Here are some examples of how this should be rated:  Example 1: "The lack of clarity might partly be caused by the fact that many RCTs were carried out on small samples and had a high risk of bias without data integrity checks, that there was a large heterogeneity in participant characteristics both within trials and between trials, and that the method and timing of scratching varied widely between studies (Li et al., 2019; Lensen et al., 2021). This has complicated conventional meta-analyses, leading to inconclusive results (Potdar et al., 2012; Nastri et al., 2015; Panagiotopoulou et al., 2015). To overcome these problems, we performed an individual participant data meta-analysis (IPD-MA) on the topic."  This study would have a rationale as "Overall treatment effect" as well as "Subgroups". They have been sufficiently clear that there are issues with conventional meta-analysis that would impact the estimation of an overall treatment effect (some reading between the lines needed, but the meaning remains sufficiently clear) and that IPD is needed to overcome this.  Example 2: "To systematically asses which is the optimal agent, we performed an individual participant data meta-analysis (IPDMA) in which we compared X with Y with respect to outcomes in XX and YY. This study has several strengths: by performing an IPDMA, we could investigate a relatively large population and this allowed for more power in subgroup analyses.".  This is not sufficiently clear. There is lacking information on needing the IPD to estimate an overall treatment effect. This is on the cusp of being a rationale of "Subgroups", but not quite sufficient. |
| IPD rationale (free-text) | Copy and paste the text where they mention their rationale, often contained in the introduction section but occasionally in the methods. |
| Protocol  available (excludes PROSPERO record)? | PROSPERO does not count here (already extracted in part 1 of this project).  If a protocol has been published, this supersedes attached to Prospero, this in turn supersedes attachment (in case the protocol is available from multiple sources).  A protocol can be:   - Published - Open Science Framework or similar - Pre-print - Protocol attached to Appendix - Protocol attached to PROSPERO record (but this needs to be a separate document, check PROSPERO record if there actually is a protocol attached, if they mention this)   This must be explicitly mentioned in the manuscript text; do not search the internet for a protocol if it has not been mentioned. |
| Statistical analysis  plan (SAP) available? | As above, this should be explicitly mentioned. Do not search the internet for a SAP. |
| Transparency 1:  Analysis model  specified? | We are looking for the equation – usually this will be in the SAP or appendix if provided at all. Scan quickly to see if you can see a formula, given in mathematical symbols or with words. If the equation is in word form, it needs to be explicit, meaning that it can be replicated without additional assumptions.  Example: Where y is a continuous outcome, z is a (set of) prognostic factors and x is the treatment allocation. y,z and x may be replaced by words that are study-specific (e.g. x=treat and z=age, say)  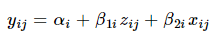 |
| Transparency 2:  Statistical code  provided? | Scroll through Appendices, skim Methods section to identify if statistical code has been mentioned.  Note that we are treating No and Not reported as the same here. |
| Which reporting guidelines followed? | Here PRISMA refers to PRISMA 2020. If PRISMA 2009 has been followed, please indicate this.  If multiple guidelines are followed, please use the Other (free text) to specify the options. |
| Risk of bias (ROB)  assessment reported? |  |
| ROB Method | Note that looking at the citation here is insufficient, since many cite the Cochrane handbook but then use ROB1 instead of ROB2. Instead, look at which tool the reported domains correspond to.  **ROB 1:** Bias is assessed as a judgement (high, low, or unclear) for individual elements from five domains of bias (selection, performance, attrition, reporting, and other).  **ROB2:** (1) bias due to the randomization process, (2) deviation from intended intervention, (3) missing outcome data, (4) measurement of outcomes, (5) selection of the reported result.  Ratings: (1) Low risk of bias; (2) Some concerns; and (3) High risk of bias.  If a study used a bespoke ROB tool that was not based on solely ROB1 or ROB2, then this would be “Other”. If a tool has been adapted directly from ROB1 or ROB2 then this would be “Adapted Cochrane” |
| Adapted Cochrane or  other tool details |  |
| IPD used to inform  RoB assessment? | Example:  “The assessments were based on information from trial protocols and manuscripts, information from trialists, and **direct checks of the individual participant data**. The direct checks were used to explore patterns of treatment allocation, the balance of baseline characteristics by treatment group, the degree of missing outcome data, how outcomes were measured, and the balance of follow-up.”  Note, the first sentence is enough to say that IPD had been used to inform RoB assessment.  Difference between not stated and no:  "Not stated" if they have a very sparse ROB section, and "No" if they detail their ROB approach and it is clear that IPD is not a part of this. |
| Data checking process reported? | Are there any reports on how data are assessed? |
| Trustworthiness checks explicitly mentioned? | This refers to checks for questionable research practices, misconduct, falsification and fabrication.  Data quality checks alone do not count (note in PRISMA-IPD data checking is labelled as integrity, so this may be confusing).  Note that checking integrity of randomisation would count as trustworthiness checks, but simply saying “checking integrity of the data” would not.  Examples for application of trustworthiness checks include:  ‘We did comprehensive, prespecified data quality and integrity checks, including items such as retraction notices, ethics approval, implausible values, and randomisation (appendix pp 23–25).’ |
| Report data checking and trustworthiness checks (if any) | Add the free text of the mentions of data checking/trustworthiness checks. This will often be in both the methods and results. |
| Was certainty of evidence assessed? |  |
| Method of certainty of  evidence assessment stated?  (If 'Other’, override w. text) |  |
| Analysis  strategy |  |
| Meta-analysis estimand reported? | Control + F search the paper, protocol and SAP  The word ‘estimand’ needs to be explicitly mentioned. |
| Analysis Type | **Frequentist analysis**  Look for confidence intervals and p-values. This is a clear indication of Frequentist analysis.  **Bayesian analysis**  Look for credible intervals, mention of priors (potentially with a distribution – or words such as weakly informative prior/non-informative prior), probability statements (e.g. 87% probability of benefit) and posterior estimates. Also, Bayesian analysis may mention using Markov Chain Monte Carlo simulation to estimate distributions.  Software packages can also tell which analyses have been performed. For example JAGS, BUGS/WinBUGS and STAN indicate Bayesian, whereas ipdmetan or metan would indicate Frequentist. |
| Number of reported  outcomes (Primary) | Count how many primary outcomes/ endpoints are listed. Usually this will be 1.  Composite counts as 1 (unless the components of the composite also analysed as primary outcomes, which should be rare)  Occasionally you may have co-primary outcomes (if there are two then this would be two primary outcomes).  Occasionally (more common with time-to-event outcomes in cancer IPD-MAs) you may have one primary outcome for main effects and one primary outcome for the subgroup effects – in this case as only one outcome is specified for each independent analysis this would still count as 1.  If no primary outcome has been specified, type: ‘not specified’ |
| Number of reported  outcomes (Secondary or other) | Count number of secondary and other outcomes. If this is unclear from methods, it may be worthwhile to look at methods as well as results table.  If there are composites plus their components, count both the overall component plus each of their components.  Example: *The secondary outcomes were ischaemic major adverse cardiovascular events (ischaemic stroke, systemic arterial embolism, pulmonary embolism, or myocardial infarction)* 🡪 this would be 5 outcomes, since both the overall category as well as the individual components were analysed as secondary outcomes  Outcomes reported and analysed at multiple timepoints (unless the only analysis is under a repeated measures framework) should be treated as separate outcomes for the purposes of counting the number. |
| Was IPD/AD combined? |  |
| Main analysis  approach | Note this is for their primary analysis (main approach only)  This information is sometimes hard to determine. See the following list of assumptions that may help to categorise:  **One-stage**   - One-step - Random slope on/for treatment - Random intercept - Mixed-effects model (or some other multilevel model described)   *If there is no mention of the two-stage options below then we can assume that the following use a one-stage approach:*   - Analysis done to entire dataset (without specifying ipdmetan or similar as software) - Stratified by/for trial - Adjusted by/for trial   **Two-stage**   - Two-step - Inferred from software used, e.g. metan,ipdmetan (Stata) or metafor (R) - Analysis within each trial then pooled across trials (or similar described) - Figures clearly show a forest plot with a two-stage approach used (ideally with trial weights) and no one-stage approach mentioned |
| Main analysis  model | Note this is for their primary analysis (main model only)  This information is sometimes hard to determine. See the following list of assumptions that may help to categorise:  **Random-effect model**   - Random slope on/for treatment - Random effect on/for treatment - Random effects model   **Fixed-effect model**   - Fixed-effect / common-effect model   *If there is no mention of the random-effects model options above then we can assume that the following are fixed-effect models:*   - Random intercept - Random effect on/for study/trial - Stratified by/for study/trial - Adjusted by/for study/trial   Note we have also asked you to copy across the text where they describe their model/approach in the field before. |
| Copy/paste main analysis approach here | Copy the text that describes their model and approach |
| Sensitivity analysis  reported? | Are any analyses explicitly labelled as sensitivity analyses (yes/ no). Use Control+F to find this. |
| Post-hoc or exploratory analysis reported? | Are any analyses explicitly labelled as post-hoc or exploratory analyses (yes/ no). Use Control+F to find this. |
| How was  heterogeneity  reported/assessed?  (Choose all that apply) | They either need to mention the statistic in the methods or report it in the results (ideally they do both). |
| Adjustment for  prognostic factors for primary analysis of main effects? | This is for their primary analysis (main approach/model only) |
| Sample size/  power calculation? | We are looking for a specific power calculation or to give the power for a specific number of patients/events. Mention of power/sample size without this would not count (e.g. “Meta-analysis can provide greater power and precision” etc.).  Look in the methods, and control+F search in the protocol/SAP if available. |
| Trial-level  subgroup analysis or meta-regression? | This is referring to trial-level factors, e.g. Blinding status, publication year, region, dose (or trials are allowed to give different doses of treatment), specific treatment (if the meta-analysis looks at a class of treatments as its primary comparison) etc.  Search for subgroup analysis and meta-regression. Note this must be a trial-level factor and not participant-level (see below for participant-level questions), so entire trials should be in one subgroup, and they should never be split across different subgroups. You may need to look at the analysis they have performed in results section/figures/tables for cases where it is hard to determine whether this is trial-level or participant level. |
| Participant-level subgroup analysis/ effect-modification analysis (Y N/not stated) | This is referring to factors that may differ at the participant-level, and that do vary in at least one trial (although often it will be in most trials). Examples would be things like Age, Sex, Disease severity (although occasionally some of these may be trial-level). Note that this can also be a continuous covariate and a categorical covariate.  You may need to look at the analysis they have performed in results section/figures/tables for cases where it is hard to determine whether this is trial-level or participant level. |
| Participant-level  subgroup analysis/effect modification analysis:  Pre-specified? | This can either be describing analyses as ‘pre-specified’ in the publication, or if the analyses are stated in a published/ time-stamped protocol.  The option “Both” may be if they have some pre-specified analyses but also some that were exploratory/post-hoc |
| Participant-level  subgroup analysis/effect modification analysis:  Hypothesis given? | We are looking for a specific hypothesis. E.g. treatment effect is more beneficial for younger ages (or similar) - might expect this in the SAP. |
| Participant-level  subgroup analysis/effect modification analysis:  How many outcomes used? | Count how many outcomes they have presented participant-level analyses for. |
| Participant-level  subgroup analysis/effect modification analysis: What outcome used? | Which of the outcomes were these analyses on? Just the primary outcome? Just the secondary outcome or multiple secondary outcomes? Or on both the primary and some secondary outcomes?  If there is no distinction between primary and secondary outcomes, then make a note of this. |
| Participant-level  subgroup analysis:  Handling of within-and  across-trial information | From IPD Handbook:   - two-stage IPD approach to estimating treatment-covariate interactions avoids aggregation bias by estimating treatment-covariate interactions in each trial separately, and then synthesising them in the second stage. This ensures that only within-trial information is used. - one-stage IPD meta-analysis to the estimation of treatment-covariate interactions must ensure that within-trial and across-trial information are separated out, by either (i) stratifying all nuisance parameters by trial, or (ii) centering the covariate by its mean and the mean covariate value to explain between-trial heterogeneity.   Example for appropriate handling/ separated out: Participant-level subgroup analyses were performed by examination of within-trial treatment-by-covariate interactions, avoiding trial-level aggregation bias by centring covariates by trial.  Note that you can often infer from the figures what they have done, especially if they use two-stage. You would be looking for a meta-analysis of interaction estimates. If unsure, then mark as unclear. This field will be checked as it is challenging to extract. |
| Approach to missing  data strategy for primary outcome analysis described? | We only care about this for the primary outcome and the analysis of main effects. Have they mentioned a strategy to deal with missing data (covariate data and/or outcome data)? |
| Type of approach to  missing data strategy | Which approach do they specify, or do they carry out multiple approaches, with one as a sensitivity analysis? |

### Supplementary File 6: Worksheet for Part 3: Consensus process to derive recommendations

**Consensus Meeting: State of play in IPD meta-analyses of RCTs**

**What will happen?**

Please take notes into your own document as you brainstorm. We will ask you to share this document after the meeting.

1. **Results interpretation.** Short brainstorms for each table, then 1 round where each person highlights 1 point from their brainstorming each, without discussion. You can pass if all your points have been taken. The facilitator records it. (6 sections)
2. **Consensus process.**
   a. There are two questions to consider, with a different amount of time allocated to each. These are at the end of the document.
   b. Each person silently thinks of solutions or ideas that come to mind when considering each question and writes down as many as possible in a set period of time. Each person states aloud one idea. The facilitator records it.
      i. No discussion is allowed, not even questions for clarification.
      ii. Ideas given do not need to be from each person’s written lists. Indeed, as time goes on, many ideas will not be found on their original lists.
      iii. You may "pass" your turn, but then can add an idea on a subsequent turn.
      iv. Continue around the group until everyone has passed or until we run out of time.
   c. Discuss each idea in turn. Wording may be changed only when the idea’s originator agrees. Ideas may be removed from the list only by unanimous agreement or when there are duplicates. Discussion may clarify meaning, explain logic or analysis, raise and answer questions, or state agreement or disagreement. The group may also combine ideas into categories.
   d. To help with prioritisation, each member will state the ideas that they feel are most important for each question.

**Section 1**

**Part 1: Results from systematic review**

The number of IPD meta-analyses of RCTs conducted as part of a systematic review has increased over time, which has now resulted in approximately 60 published per year.

*see Figure 1*

The main clinical areas studied were diseases of the circulatory system (19%), neoplasms (18%) and mental, behavioural or neurodevelopmental disorders (12%). The median number of trials that provided IPD was 7 [4, 14], with the median number of participants with IPD included 2096 [787, 5948]. The majority had corresponding author based in Europe (70%).

*see Table 1*

**Part 2: Current practice in the conduct, analysis and reporting of IPD meta-analyses**

For Part 2, we limited the detailed extraction to just pairwise IPD meta-analyses that were published across a two-year period (26 February 2022 to 26 February 2024). This resulted in 100 unique studies. The remainder of this results document describes these 100 IPD meta-analyses.

*see Supplementary Table 1*

**Prompts for section 1:**

- **Which of these results are most noteworthy?**
- **Are there key take aways, lessons or recommendations?**

***Aim 2: To describe data retrieval rates and approaches to handling studies without IPD and evaluation of data availability bias (e.g. inclusion of unpublished trials, combination of IPD with AD)***

**Section 2**

*see Supplementary Table 2*

**Prompts for section 2:**

- **Which of these results are most noteworthy?**
- **Are there key take aways, lessons or recommendations?**

***Aim 3: To evaluate the design, conduct and reporting of IPD-MAs.***

**Section 3**

*see Supplementary Table 3*

**Prompts for section 3:**

- **Which of these results are most noteworthy?**
- **Are there key take aways, lessons or recommendations?**

**Section 4**

*see Supplementary Table 4*

**Prompts for section 4:**

- **Which of these results are most noteworthy?**
- **Are there key take aways, lessons or recommendations?**

***Aim 4: To summarise the statistical analysis strategies (and their prevalence) employed by current IPD-MAs of RCTs***

**Section 5**

see Supplementary Table 5

**Prompts for section 5:**

- **Which of these results are most noteworthy?**
- **Are there key take aways, lessons or recommendations?**

**Section 6**

*see Supplementary Table 6*

**Prompts for section 6:**

- **Which of these results are most noteworthy?**
- **Are there key take aways, lessons or recommendations?**

**Prompts for consensus process**

*Suggested timings for each part are indicated in brackets*

**Question 1**

**What do these results tell us about the current state of play in IPD meta-analysis?**

- Brainstorm (2 mins)
- Round robin – everyone states one point in turn, we keep going around until no one has anything additional to say or time runs out, facilitator records it (5 mins)
- Discuss each point, combine, summarise, remove duplicates (8 mins)
- Agree on the most important points (2 mins)

**Question 2**

**Where do we need to develop new guidance or provide better access to existing methods?**

- Brainstorm (3 mins)
- Round robin – everyone states one point in turn, we keep going around until no one has anything additional to say or time runs out, facilitator records it (7 mins)
- Discuss each point, combine, summarise, remove duplicates (15 mins)
- Agree on the most important points (3 mins)

### Supplementary Tables and Figures

#### Supplementary Figure 1: PRISMA Flow diagram

Studies from databases/registers **(n = 5004)**

**Identification**

References removed **(n = 416)**

Duplicates identified manually (n = 55)

Duplicates identified by Covidence (n = 361)

Studies excluded **(n = 566)**

IPD-MA of non-RCTs (n = 240)

Conference abstract (n = 120)

No systematic search (n = 102)

Not a MA (e.g. editorial, letter) (n = 29)

No IPD (n = 33)

Can't find text (n = 17)

Wrong study design (e.g. surrogacy) (n=13)

Protocol only (n = 4)

Methodological paper (n = 3)

Not published in English (n = 2)

Only single arms from RCTs used (n = 2)

Duplicate (n=1)

Studies assessed for eligibility **(n = 848)**

Studies screened **(n = 4588)**

Studies excluded **(n = 3740)**

**Screening**

Eligible studies from searches **(n = 282)**

Studies from Wang et al. [8] **(n = 323)**

**Included**

Studies included in review **(n = 605)**

#### Supplementary Table 1: General characteristics

| **Characteristic** | **Total (n=100)** |
| --- | --- |
| **Medical field** |  |
| Mental, behavioural or neurodevelopmental disorders | 19 (19%) |
| Diseases of the circulatory system | 18 (18%) |
| Neoplasms | 11 (11%) |
| Pregnancy, childbirth or the puerperium | 10 (10%) |
| Musculoskeletal system or connective tissue | 8 (8%) |
| Certain infectious or parasitic diseases | 8 (8%) |
| Endocrine, nutritional or metabolic diseases | 5 (5%) |
| Diseases of the digestive system | 3 (3%) |
| Other | 18 (18%) |
| **Cochrane Review** |  |
| Yes | 3 (3%) |
| No | 97 (97%) |
| **Update to a previous review** |  |
| Yes | 12 (12%) |
| No | 86 (86%) |
| Unclear | 2 (2%) |
| **Type of review** |  |
| Retrospective | 87 (87%) |
| Prospective | 9 (9%) |
| Nested prospective | 4 (4%) |
| **Geographical location of corresponding author** |  |
| Australia | 18 (18%) |
| UK | 17 (17%) |
| The Netherlands | 17 (17%) |
| USA | 14 (14%) |
| Germany | 9 (9%) |
| Switzerland | 5 (5%) |
| Spain | 3 (3%) |
| Japan | 3 (3%) |
| France | 2 (2%) |
| Other | 12 (12%) |
| **Funding status** |  |
| Public only | 50 (50%) |
| Charity/Philanthropic only | 7 (7%) |
| Industry only | 5 (5%) |
| Public & Charity/Philanthropic | 13 (13%) |
| Public & Industry | 3 (3%) |
| Industry & Charity/ Philanthropic | 1 (1%) |
| Public & Charity/Philanthropic & Industry | 4 (4%) |
| None/not stated | 17 (17%) |

Data are n (%)

#### Supplementary Table 2: Data sources and retrieval

| **Characteristic** | **Total (n=100)** |
| --- | --- |
| **Trials** |  |
| ***Identified as eligible*** |  |
| Median [25^th^, 75^th^] | 9 [4, 17] |
| Min, Max | 2, 122 |
| Unclear | 1 (1%) |
| ***IPD obtained from*** |  |
| Median [25^th^, 75^th^] | 6 [4, 10] |
| Min, Max | 2, 48 |
| ***Excluding 1 with Unclear data, IPD obtained from*** |  |
| Median [25^th^, 75^th^] | 6 [4, 10] |
| Min, Max | 2, 48 |
| ***Proportion obtained**** |  |
| Median [25^th^, 75^th^] | 85.7% [50.0%, 100%] |
| Min, Max | 19.0%, 100% |
| **Participants** |  |
| ***Identified as eligible*** |  |
| Median [25^th^, 75^th^] | 3178 [1087, 9519] |
| Min, Max | 131, 154664 |
| Unclear | 17 (17%) |
| ***IPD obtained from*** |  |
| Median [25^th^, 75^th^] | 2174.5 [669.5, 6972.5] |
| Min, Max | 131, 154664 |
| ***Excluding 17 with Unclear data, IPD obtained from*** |  |
| Median [25^th^, 75^th^] | 2313 [771, 7529] |
| Min, Max | 131, 154664 |
| ***Proportion obtained**** |  |
| Median [25^th^, 75^th^] | 99.2% [75.6%, 100%] |
| Min, Max | 8.9%, 100% |
| **Data obtained from** |  |
| Investigators only | 82 (82%) |
| Repositories only | 2 (2%) |
| Investigators and repositories | 7 (7%) |
| Investigators and supplied as a supplement to journal publication | 1 (1%) |
| Other | 2 (2%) |
| Not stated | 6 (6%) |
| ***If data obtained from repositories, was this downloadable*** |  |
| All downloadable | 2 (22%) |
| None downloadable | 1 (11%) |
| Not stated | 6 (67%) |
| **Was aggregate data extracted** |  |
| Yes | 33 (33%) |
| ***If Yes, number of extracted trials*** |  |
| Median [25^th^, 75^th^] | 7 [2, 15] |
| Min, Max | 1, 116 |
| Median proportion of aggregate data as total* | 50% [25%, 60%] |
| ***If Yes, number of extracted participants*** |  |
| Median [25^th^, 75^th^] | 1355 [274, 2927] |
| Min, Max | 64, 64439 |
| Median proportion of aggregate data as total* | 41.4% [10.2%, 53.4%] |
| Unclear | 3 (9%) |
| **Searching trial registries** |  |
| Yes | 55 (55%) |
| No | 39 (39%) |
| Not stated | 6 (6%) |
| **Unpublished trials obtained** |  |
| Yes | 15 (15%) |
| No/not stated | 63 (63%) |
| NA – all eligible trials published | 22 (22%) |
| ***If Yes, number of unpublished trials*** |  |
| Median | 1.5 [1, 4.5] |
| Min, Max | 1, 11 |
| Unclear | 3 (20%) |
| ***If Yes, number of participants from unpublished trials*** |  |
| Median | 158.5 [88, 1032] |
| Min, Max | 15, 9938 |
| Unclear | 5 (33%) |

Data are n (%) unless stated
*Proportion obtained are from a within-study comparison only

#### Supplementary Table 3: Design, conduct and reporting: rationale and transparency

| **Characteristic** | **Total (n=100)** |
| --- | --- |
| **Clear rationale for IPD meta-analysis** |  |
| No | 38 (38%) |
| Yes | 62 (62%) |
| ***If Yes, Rationale**** |  |
| Subgroups | 45 (73%) |
| Overall treatment effect | 42 (68%) |
| Harmonisation | 7 (11%) |
| Obtain longer term follow-up | 6 (10%) |
| Obtain unpublished data | 4 (6%) |
| **Registered** |  |
| Yes | 68 (68%) |
| No/not stated | 32 (32%) |
| **Protocol available** |  |
| Available and published in a peer-reviewed journal | 22 (22%) |
| Available and attached to appendix | 6 (6%) |
| Available on a pre-print server | 4 (4%) |
| Available and attached to PROSPERO registration | 1 (1%) |
| Not available/mentioned | 67 (67%) |
| **Statistical analysis plan available** |  |
| Yes | 29 (29%) |
| No | 71 (71%) |
| **Reporting guideline*** |  |
| PRISMA-IPD | 66 (66%) |
| PRISMA 2020 | 15 (15%) |
| PRISMA 2009 | 1 (1%) |
| STROBE | 1 (1%) |
| MECIR | 1 (1%) |
| No/not stated | 17 (17%) |
| **Statistical transparency** |  |
| Statistical model specified | 7 (7%) |
| Analysis code provided | 8 (8%) |

Data are n (%)
*Not mutually exclusive

#### Supplementary Table 4: Design, conduct and reporting: data quality, bias, certainty of evidence

| **Characteristic** | **Total (n=100)** |
| --- | --- |
| **Risk of bias assessment** |  |
| No | 18 (18%) |
| Yes | 82 (82%) |
| ***If Yes, Risk of Bias method*** |  |
| Cochrane Risk of Bias tool 1 | 28 (34%) |
| Cochrane Risk of Bias tool 2 | 46 (56%) |
| Adapted Cochrane | 3 (4%) |
| Other | 5 (6%) |
| ***If Yes, was IPD used to inform Risk of Bias assessment?*** |  |
| Yes | 6 (7%) |
| No/Not stated | 76 (93%) |
| **Data checking process reported** |  |
| Yes | 68 (68%) |
| No | 32 (32%) |
| **Trustworthiness checks explicitly reported** |  |
| Yes | 15 (15%) |
| No | 78 (78%) |
| Unclear | 7 (7%) |
| **Certainty of evidence assessed** |  |
| Yes | 27 (27%) |
| ***If Yes, method*** |  |
| GRADE | 26 (96%) |
| Not stated | 1 (4%) |

Data are n (%)

#### Supplementary Table 5: Statistical analysis strategies: Main/overall effects

| **Characteristic** | **Total (n=100)** |
| --- | --- |
| **Estimand reported** |  |
| Yes | 1 (1%) |
| No | 99 (99%) |
| **Sample size/power calculation a priori** |  |
| Yes | 4 (4%) |
| No/not stated | 96 (96%) |
| **Analysis type** |  |
| Frequentist | 93 (93%) |
| Bayesian | 5 (5%) |
| Both | 2 (2%) |
| **Number of outcomes specified as primary** |  |
| Median [25^th^, 75^th^] | 1 [1, 2] |
| Min, Max | 0, 25 |
| Unclear | 2 (2%) |
| **Number of other outcomes** |  |
| Median [25^th^, 75^th^] | 5 [2, 9] |
| Min, Max | 0, 29 |
| Unclear | 2 (2%) |
| **Was IPD and AD combined?** |  |
| Yes, main analysis | 9 (9%) |
| Yes, sensitivity analysis | 16 (16%) |
| No/not stated | 50 (50%) |
| NA, IPD from all eligible trials obtained | 25 (25%) |
| **Main analysis approach** |  |
| One-stage | 52 (52%) |
| Two-stage | 35 (35%) |
| Both, without specifying a main approach | 9 (9%) |
| Unclear | 4 (4%) |
| **Main analysis model** |  |
| Common/fixed effect(s) | 30 (30%) |
| Random effects | 50 (50%) |
| Both, without specifying a main model | 7 (7%) |
| Unclear | 13 (13%) |
| **Main analysis model adjusted for covariates/prognostic factors?** |  |
| Yes | 50 (50%) |
| No/Unclear | 50 (50%) |
| **Missing data approach for main analysis described** |  |
| Yes | 55 (55%) |
| No | 45 (45%) |
| ***If Yes, approach used*** |  |
| Complete case analysis | 17 (31%) |
| An imputation method | 27 (49%) |
| Both, with one as a sensitivity analysis | 10 (18%) |
| Other | 1 (1%) |
| **How was statistical heterogeneity identified/measured*** |  |
| Cochran’s Q test | 17 (17%) |
| Visual inspection of forest plots | 4 (4%) |
| I^2^ statistic | 59 (59%) |
| Tau^2^ | 26 (26%) |
| Prediction interval | 1 (1%) |
| Breslow-day test of homogeneity | 1 (1%) |
| Not reported | 24 (24%) |

Data are n (%) unless stated
*Not mutually exclusive

#### Supplementary Table 6: Statistical analysis strategies: Subgroup/interaction effects

| **Characteristic** | **Total (n=100)** |
| --- | --- |
| **Trial-level subgroup analysis and/or trial-level meta regression reported** |  |
| Yes | 24 (24%) |
| No/not stated | 76 (76%) |
| **Participant-level subgroup/interaction analysis reported** |  |
| Yes | 85 (85%) |
| No/not stated | 15 (15%) |
| ***If Yes, pre-specified?*** |  |
| Pre-specified only | 45 (53%) |
| Post-hoc only | 3 (4%) |
| Both | 20 (24%) |
| Unclear | 17 (20%) |
| ***If Yes, hypothesis given?*** |  |
| Yes | 11 (13%) |
| No | 74 (87%) |
| ***If Yes, number of outcomes used*** |  |
| Median [25^th^, 75^th^] | 2 [1, 5] |
| Min, Max | 1, 25 |
| Unclear | 4 (5%) |
| ***If Yes, what was outcome?*** |  |
| Primary outcome(s) for main effects only | 44 (52%) |
| Secondary outcome(s) for main effects only | 2 (2%) |
| Both primary and secondary outcome(s) for main effect | 36 (42%) |
| Unclear | 3 (4%) |
| ***If Yes, within and across-trial information separated out?*** |  |
| Separated out | 35 (41%) |
| Conflated | 15 (18%) |
| Unclear | 35 (41%) |

Data are n (%) unless stated

#### Supplementary Table 7: Demographics of consensus panel members

|  | **Consensus panel (n=24)** |
| --- | --- |
| **Location of primary affiliation** |  |
| UK | 13 (54%) |
| Australia | 6 (25%) |
| Switzerland | 3 (13%) |
| Singapore | 1 (4%) |
| USA | 1 (4%) |
| **Career stage** |  |
| Early-career researcher | 8 (33%) |
| Mid-career researcher | 6 (25%) |
| Experienced researcher | 10 (42%) |
| **Years of experience in IPD-MA** |  |
| 0-4 | 6 (25%) |
| 5-9 | 5 (21%) |
| 10+ | 13 (54%) |
| **Areas of expertise*** |  |
| Undertaking IPD-MA | 24 (100%) |
| Developing IPD-MA methodology | 19 (79%) |
| Systematic reviews | 19 (79%) |
| Statistics | 11 (46%) |
| Clinical trials | 9 (38%) |
| Guideline development | 5 (21%) |
| Clinical medicine | 4 (17%) |
| Health technology assessment | 3 (13%) |
| **Main medical area(s) specialised in*** |  |
| Cancer | 9 (38%) |
| Infectious diseases | 5 (21%) |
| Neonatology | 5 (21%) |
| Mental health | 4 (17%) |
| Child health | 4 (17%) |
| Cardiovascular | 3 (13%) |
| Public health | 3 (13%) |
| Women’s health | 2 (8%) |
| Diabetes | 2 (8%) |
| Critical care | 1 (4%) |
| Musculoskeletal | 1 (4%) |
| *No specific area* | 3 (13%) |

*Not mutually exclusive
IPD-MA=Individual participant data meta-analysis
